# Sibling Variation in Phenotype and Genotype: Polygenic Trait Distributions and DNA Recombination Mapping with UK Biobank and IVF Family Data

**DOI:** 10.1101/2022.09.17.22280057

**Authors:** Louis Lello, Maximus Hsu, Erik Widen, Timothy G. Raben

## Abstract

We use UK Biobank and a unique IVF family dataset (including genotyped embryos) to investigate sibling variation in both phenotype and genotype. We compare phenotype (disease status, height, blood biomarkers) and genotype (polygenic scores, polygenic health index) distributions among siblings to those in the general population. As expected, the between-siblings standard deviation in polygenic scores is 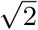 times smaller than in the general population, but variation is still significant. As previously demonstrated, this allows for substantial benefit from polygenic screening in IVF. Differences in sibling genotypes result from distinct recombination patterns in sexual reproduction. We develop a novel sibling-pair method for detection of recombination breaks via statistical discontinuities. The new method is used to construct a dataset of 1.44 million recombination events which may be useful in further study of meiosis.

## 1 Introduction

It is commonly understood that siblings are more similar to each other and their parents than to unrelated individuals in the general population. Consequently there is less variation (e.g., in height or hair color) among siblings from the same family than in a group of randomly selected, unrelated individuals. In this paper we explore these observations from the perspective of genetic similarity and DNA inheritance – see **Figure 1**. We study both realized phenotypes as well as genotypes, in particular in the form of polygenic scores (PGS). PGS are typically linear functions of thousands of genetic loci, constructed via machine learning on datasets with human genotype and phenotype association. PGS exist for a large variety of phenotypes [1–9] (such as disease condition status, BMI, height, etc.), they have numerous potential applications in public health and in clinical settings [5, 10–30], but important barriers still exist to widespread clinical adoption [31–38].

**Figure 1:**
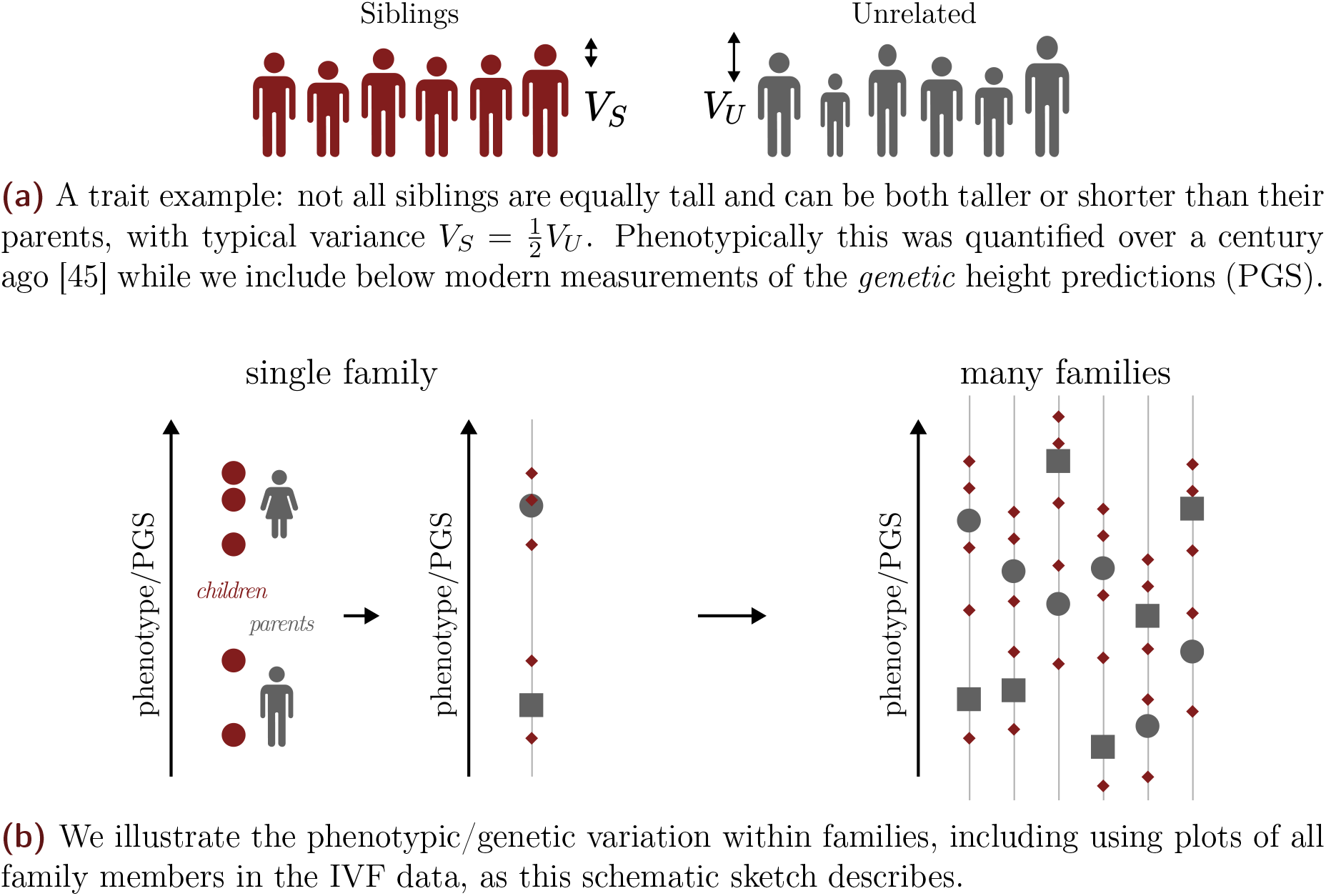
Trait and genetic variation among siblings is smaller than in the general population, but still significant. Population genetics theory implies that the standard deviation among siblings is 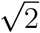 smaller than for the general population. We demonstrate this empirically using UKB and IVF clinic data, and illustrate that variation in important traits and disease risks among siblings is substantial.

Full siblings are identical at approximately half of all DNA loci, and each receives approximately half of their DNA from each parent. This has the following implications for complex traits that are roughly additive in genetic architecture:

1. The sibling distribution of (additive) polygenic scores (PGS) for such traits is centered at the parental midpoint
2. The sibling genetic variance is roughly half that found in the general population (i.e., of individuals who are not closely related to each other). The probability distribution of sibling polygenic scores has standard deviation 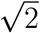 times smaller than in the general population [39].

Whether observed phenotypes also exhibit these properties depends further on the nature of environmental influences, and interactions between genotype and environment [40]. If environmental influences are themselves roughly additive, and mostly independent of genotype (and of parental background), we expect that sibling phenotypes are also normally distributed, roughly about the parental midpoint, with reduced variance compared to that found in the general population. The degree of reduction in variance depends on the heritability of the trait in question.

We examine the properties discussed above from an empirical (or descriptive) perspective, using sibling and family data from UK Biobank (UKB) and from in-vitro fertilization (IVF) clinics. In the latter case routine embryo biopsies taken for genetic screening purposes have been genotyped by the company Genomic Prediction (GP), using an Affymetrix array similar to the one used by UKB. Specifically, we examine variance in and correlation between sibling genotypes and phenotypes across a number of complex traits, developing evidence for observations 1. and 2. above.

A surprising objection to the use of PGS in the context of IVF is that the variation within families (specifically between siblings) is negligible and therefore it has little utility as a screening mechanism [41, 42]. For example, in [42] the authors write

> “It is also important to note that all embryos produced by a couple are genetically related and share on average 50% of SNPs [single nucleotide polymorphism]. One must conclude that owing to inherent limitations of the PRS-ES [polygenic risk score for embryo screening] models and limited variation in the genetic makeup of embryos produced by a couple, the clinical utility of PRS-ES is almost certainly diminutively small.”

As we illustrate below, the genetic variation among embryos (siblings) is quite substantial: it is common to find, within a specific family, embryos that are both high and normal risk for a specific condition, or even in overall Genetic Health Index score, a composite predictor of multiple disease risks with the purpose of capturing general health [43]. That PGS have strong performance among siblings (comparable to, but slightly weaker than in unrelated pairs) has already been demonstrated and repeatedly verified in previous work [27, 43, 44]. In section 4.2 we review standard results from population genetics which are relevant to sibling genetic variation. These results can be tested empirically in the modern era of large genomic biobank data and in light of PGS.

Family data can be further used to explore the specific genotype differences between siblings. These arise through the randomness of the recombination process that produces the sperm and egg genotypes in reproduction. We develop a new *sibling-pair method* for detecting recombination events by comparing the genotypes of two siblings. Large biobanks such as UKB have tens of thousands of sibling pairs, which allow high-statistics mapping of recombination rates in sexual reproduction.

The sibling-pair method makes use of the following observation (see section 4.3 for more details). Two siblings, sib1 and sib2, will have identical DNA at almost every locus in certain regions (i.e., identical by descent or IBD) which, in total, amount to roughly one fourth of their entire genome. The boundaries of these regions are easy to detect: crossing such a boundary causes the average pairwise similarity to drop from nearly 100 percent to a much lower value. We can search for adjacent blocks of SNPs with very different pairwise (sib1 vs sib2) similarity. The boundary separating these blocks is a recombination event, and the distribution of these boundaries characterizes the recombination rate as a function of position on each chromosome.

Note that while these regions where sib1 and sib2 have exactly the same DNA sequence are particularly easy to detect and analyze (i.e., to find where they begin and end), nevertheless the recombination events themselves, which take place in the meiosis that produces a specific sperm or egg cell, are obviously independent of each other (sperm vs egg, or sib1 vs sib2). Hence the location of each boundary of these special regions is an unbiased and independent draw from the probability distribution which characterizes recombination. Cataloging the location of these boundaries therefore allows us to estimate this probability distribution – i.e., the recombination rate (in Morgans) as a function of position on each chromosome. Because this method of recombination mapping only requires the genotypes of two siblings, it can be applied to very large existing datasets.

As a check on the method, we test it using datasets in which we have access to parental as well as sibling genotypes. IVF family data is especially useful here because the number of embryos is typically larger than the number of siblings in a family, and the number of pairwise analyses one can use for testing is large. By specifically looking at SNPs where the mother and father are homozygous and heterozygous (or vice versa), we can easily track recombination events which switch the chromatid region that is passed on to the child. We have used smaller datasets with both parent and sibling data to test and calibrate the sibling-only method described above. See section 2.4 and section 4.3 for more details.

We have developed the sibling-pair method for detecting recombination breaks using imputed SNP genotypes. This limits the spatial resolution of the results relative to what can be obtained using whole genomes, however it is sufficient to map out the probability distribution and to locate recombination hotspots (i.e., where the local recombination rate is much larger than the average for the chromosome). In this paper we apply the method to imputed SNP data from UKB and from an IVF family dataset.

The sibling-pair method can also be applied to whole genome data, which would result in more precise spatial resolution. We anticipate that large datasets of recombination events can be analyzed using machine learning techniques to identify the specific DNA features (e.g., motifs) which are associated with higher probability of recombination in a given region.

In this paper we use a number of different phenotypes and PGS, based on previous publications[3, 27, 43, 44], on siblings and unrelated populations. See the Supplement section 1.3 for additional information on how the phenotypes are specified and the PGS are trained. In addition to the PGS, there is a Genetic Health Index, which is a sum of 20 PGS-derived disease risks weighted by life expectancy impact of each disease. The specific index used in this work is described in [43], including details of its construction and properties. Additional information about biobanks can be found in section 4.1 and in the supplement, section 1.

## 2 Results

Modern developments such as large-scale genetic biobanks and PGS allow us to explore a number of quantities that arise in classical population genetics. We present such results in this section. In addition, we correct some misconceptions found in the literature [42] concerning within family variation of PGS/phenotype compared to the general population.

### 2.1 Overview of Methods

Some basic explanation of the notation is required before presenting the results. The methods themselves, including details about recombination, are described in section 4.

We repeatedly compare the PGS differences among unrelated individuals and siblings. Δ*U* denotes the difference, or the distribution thereof, between unrelated individuals paired randomly. That is, the PGS difference between two randomly chosen individuals. Δ*S* denotes the difference (or distribution of differences) between pairs of genetic siblings.

We extract the assortativity *m* from the covariance between the PGS (trained on phenotypes adjusted for common covariates like age and sex) of two siblings, *S*_1_, *S*_2_, and the general PGS variance *V* according to

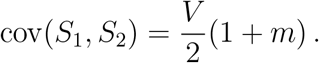

We compute the narrow-sense heritability *h*^2^ as the average of two methods: (1) the ratio between PGS variance and phenotypic variance, and (2) the square of the correlation between PGS and phenotypes.

Lastly, we also present results for a Genetic Health Index, which is a linear combination of absolute risks estimated from PGS [43]. Expressed in terms of the average lifespan reduction *l*_*d*_, average lifetime risk *ρ*_*d*_ and PGS-estimated individual risk *r*_*d*_ for each disease *d*, the Genetic Health Index is

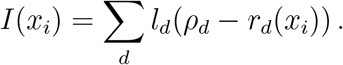

The particular index used includes risk predictors for 20 common and impactful diseases: Alzheimer’s disease, asthma, atrial fibrillation, basal cell carcinoma, breast cancer, coronary artery disease, gout, heart attack, high cholesterol, hypertension, inflammatory bowel disease, ischemic stroke, major depressive disorder, malignant melanoma, obesity, prostate cancer, schizophrenia, testicular cancer, and type I & II diabetes. See section 4.2.1 for details on the risk model, etc.

### 2.2 Sibling vs Unrelated PGS results in UK Biobank

The results discussed here provide an empirical exploration of the theoretical predictions given in section 4.2.

We identify the set of sibling pairs of self-reported European (White, in UKB terminology) ancestry within the UKB. We construct a set of randomly paired individuals: each individual is assigned to a pair with the partner chosen at random from the aggregate sibling group. We then calculate the phenotype difference between sibling pairs and random pairs with the expectation that the sibling pairs will display less variation that that of the random pairs. In **Figure 2**, we show the difference in phenotype versus the difference in PGS between sibling pairs and random pairs for height and BMI. **Figure 2** exhibits the similarity of sibling pairs in phenotype and PGS. Additionally several quantities can be extracted from sibling data. The narrow-sense heritability captured by a polygenic predictor can be estimated by taking the ratio of phenotype-to-PGS variation or by computing phenotype-to-PGS correlation (see eq. (3) and eq. (5)). We present the average of these two methods. The correlation of trait values between sibling pairs and total additive variance can be used to estimate assortivity, see eq. (6).

**Figure 2:**
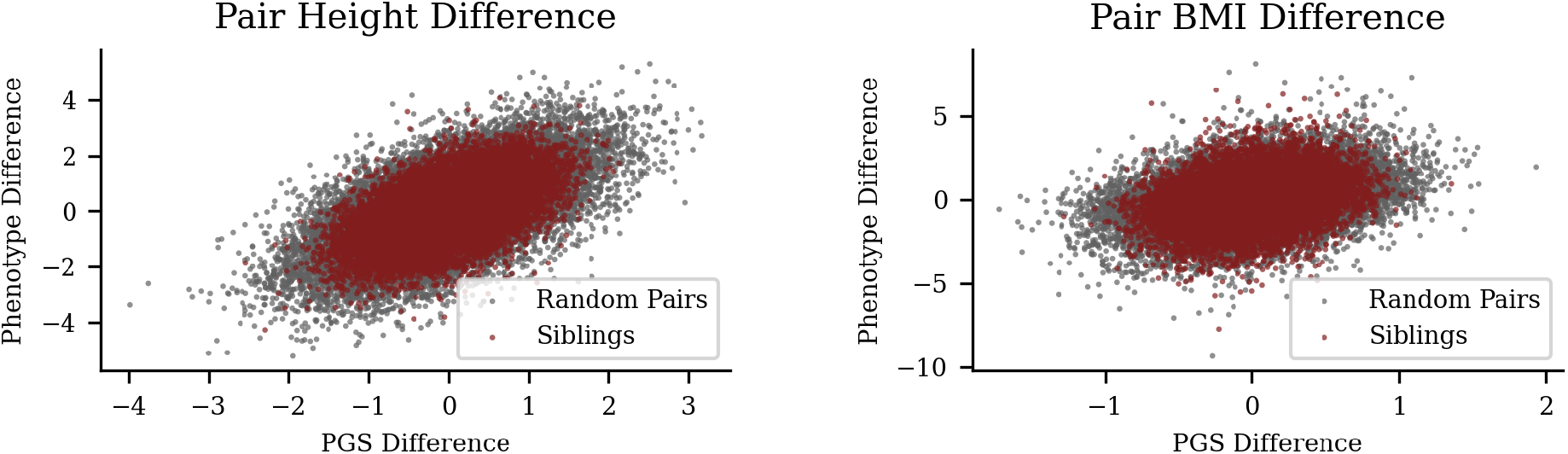
Reduced, but still significant, variation among siblings relative to general population. UKB sibling pair vs unrelated pair differences in phenotype (vertical axis) and corresponding difference in PGS (horizontal axis). The left and right panels are height and BMI respectively.

In **Table 1**, for quantitative phenotypes we list PGS and phenotype correlations between siblings, the narrow-sense heritability, and the implied assortivity. We also compare the standard deviations found in sibling (*S*) and unrelated (*U*) distributions to test whether the former are smaller by 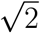.

**Table 1:** Descriptive population genetics results for sibling quantitative traits: the ratio of unrelated (*U*) vs sibling (*S*) standard deviations holds to be 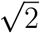. Correlation in PGS and phenotypes (adjusted by sex) between pairs of siblings, trait heritability, and assortivity *m* for quantitative traits. *σ*(Δ*U*) is the standard deviation of the PGS-difference among unrelated individuals and *σ*(Δ*S*) is the corresponding standard deviation among siblings. Computed using UKB siblings of either sex. All uncertainties, listed in parentheses, are standard errors.

| Phenotype | $N$ pairs | PGS-corr. | phen. corr. | $h^2$ | $m$ | $\sqrt{2} \frac{\sigma(\Delta S)}{\sigma(\Delta U)}$ |
| --- | --- | --- | --- | --- | --- | --- |
| Height | 21,404 | 0.559<sub>(0.006)</sub> | 0.520<sub>(0.006)</sub> | 0.385<sub>(0.004)</sub> | 0.12<sub>(0.02)</sub> | 0.939<sub>(0.006)</sub> |
| Bone Density | 10,126 | 0.501<sub>(0.009)</sub> | 0.275<sub>(0.009)</sub> | 0.16<sub>(0.01)</sub> | 0.00<sub>(0.02)</sub> | 1.010<sub>(0.010)</sub> |
| BMI | 21,064 | 0.520<sub>(0.006)</sub> | 0.287<sub>(0.006)</sub> | 0.106<sub>(0.007)</sub> | 0.04<sub>(0.02)</sub> | 0.982<sub>(0.007)</sub> |
| Fluid Int. | 4,967 | 0.55<sub>(0.01)</sub> | 0.33<sub>(0.01)</sub> | 0.071<sub>(0.007)</sub> | 0.11<sub>(0.03)</sub> | 0.94<sub>(0.010)</sub> |
| Edu Years | 21,333 | 0.591<sub>(0.006)</sub> | 0.320<sub>(0.006)</sub> | 0.070<sub>(0.007)</sub> | 0.18<sub>(0.02)</sub> | 0.907<sub>(0.006)</sub> |
| Systolic BP | 19,647 | 0.505<sub>(0.006)</sub> | 0.167<sub>(0.006)</sub> | 0.038<sub>(0.009)</sub> | 0.01<sub>(0.02)</sub> | 0.990<sub>(0.007)</sub> |
| Diastolic BP | 19,647 | 0.514<sub>(0.006)</sub> | 0.175<sub>(0.006)</sub> | 0.064<sub>(0.004)</sub> | 0.03<sub>(0.02)</sub> | 0.979<sub>(0.007)</sub> |
| HDL | 16,850 | 0.491<sub>(0.007)</sub> | 0.286<sub>(0.007)</sub> | 0.199<sub>(0.005)</sub> | -0.02<sub>(0.02)</sub> | 1.004<sub>(0.008)</sub> |
| LDL | 19,823 | 0.495<sub>(0.006)</sub> | 0.173<sub>(0.006)</sub> | 0.096<sub>(0.007)</sub> | -0.01<sub>(0.02)</sub> | 1.003<sub>(0.007)</sub> |
| Lipoprotein A | 13,383 | 0.460<sub>(0.008)</sub> | 0.358<sub>(0.008)</sub> | 0.56<sub>(0.01)</sub> | -0.08<sub>(0.02)</sub> | 1.000<sub>(0.009)</sub> |
| Total Bilirubin | 19,732 | 0.510<sub>(0.006)</sub> | 0.275<sub>(0.006)</sub> | 0.343<sub>(0.005)</sub> | 0.02<sub>(0.02)</sub> | 1.002<sub>(0.007)</sub> |
| Platelet Count | 20,521 | 0.508<sub>(0.006)</sub> | 0.278<sub>(0.007)</sub> | 0.21<sub>(0.01)</sub> | 0.02<sub>(0.02)</sub> | 0.992<sub>(0.007)</sub> |

For case-control phenotypes, we can still observe the PGS difference and correlation between siblings in PGS and also demonstrate that the variance in scores between siblings is quite substantial. In **Figure 3** we display the difference in sibling vs random pairs for PGS in Type 2 Diabetes; a corresponding figure for genetic health index (see eq. (12)) is shown in **Figure 4**. *In* ***Table 2*** *we compute PGS correlations, implied assortativity m*, and test the 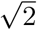 relationship between sibling (*S*) and unrelated (*U*) standard deviation in PGS. The interpretation of *m* here (naively, negative assortation on certain disease risk polygenic scores) is unclear to us and deserves further investigation.

**Figure 3:**
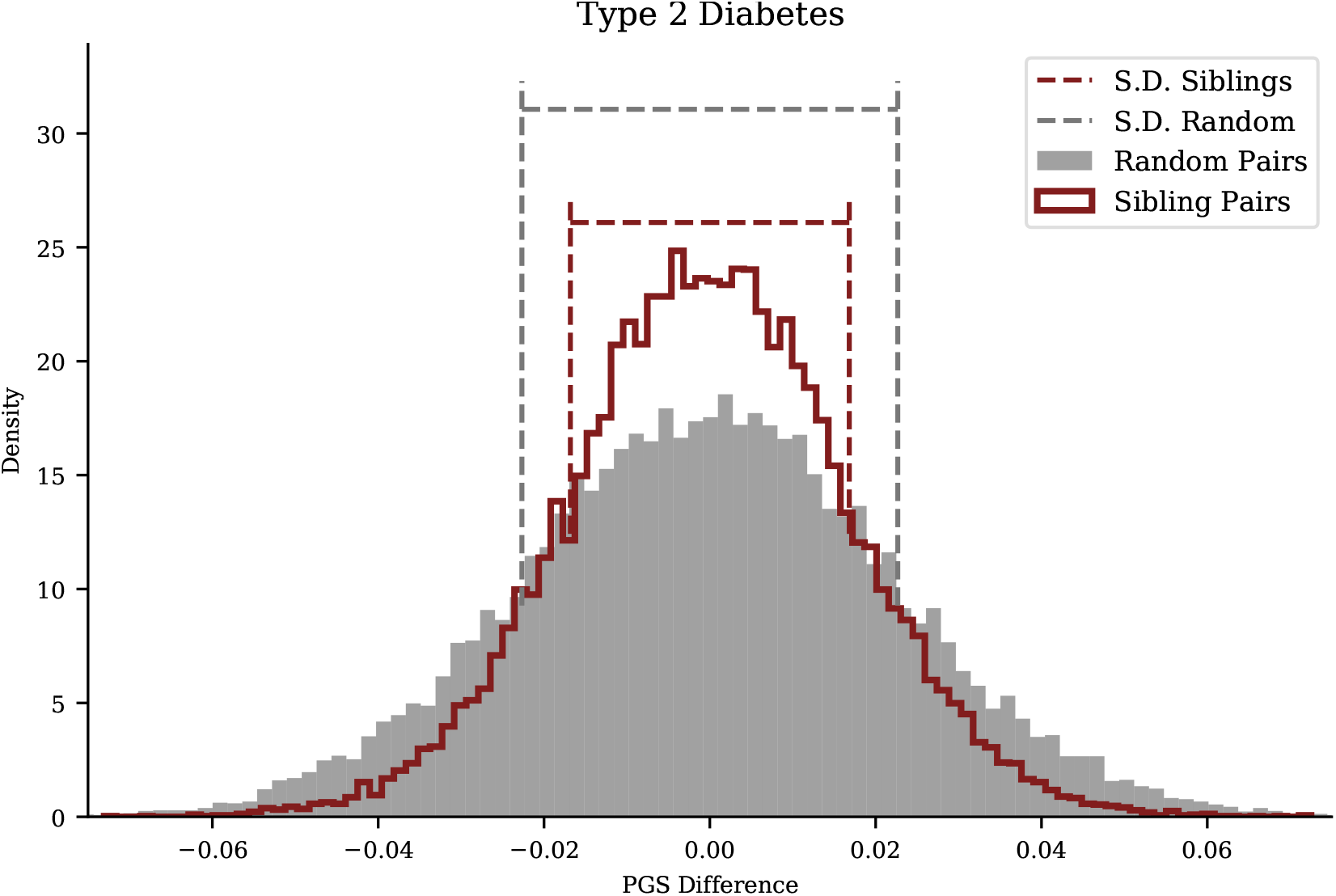
Type 2 Diabetes PGS: sibling variation smaller but still significant. Difference in PGS for Type 2 Diabetes in pairs of siblings and pairs of unrelated individuals from the general UKB population. The width of the sibling distribution is, as expected, 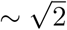 times smaller, but still significant. The dashed lines mark the standard deviations for the distributions.

**Figure 4:**
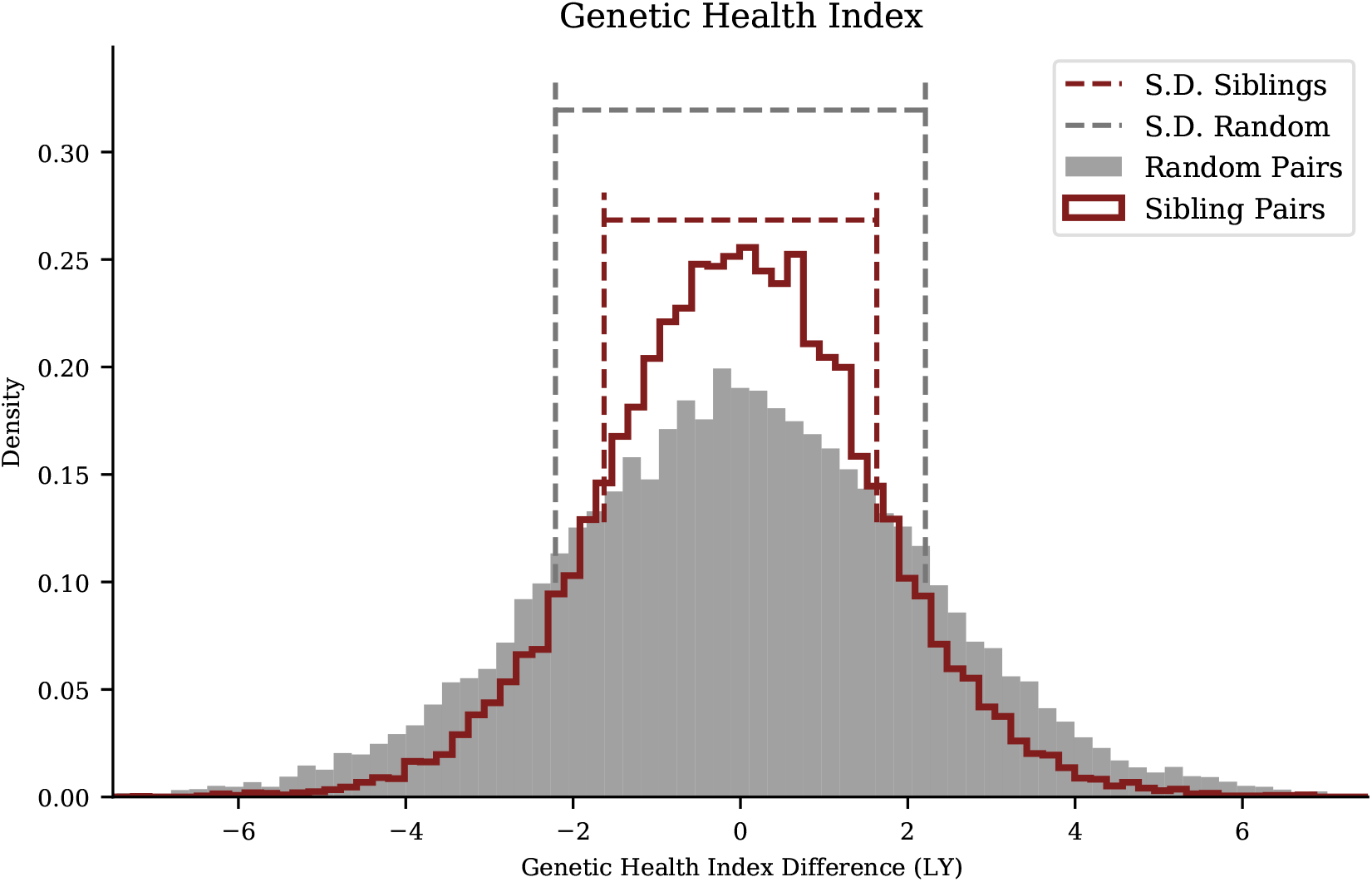
Health Index Polygenic Score: reduced, but still significant, variation among siblings relative to general population. For UKB sibling pair vs unrelated pair differences in Health Index we also find 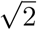 reduction in standard deviations of PGS differences for siblings, as indicated by the dashed lines.

These calculations summarize how the collection of sibling pairs compares to the collection of unrelated pairings. We now explicitly show that individual families display a level of variance comparable to what is claimed. In the prior plots and tables, we focused solely on *pairs* of (full) siblings – however, in the UK Biobank, larger family structures are present. We identify larger families by collecting all sets such that if A-B and B-C are full sibling pairs, the superset A-B-C must be a family (we filter against parent-child relationships). The number and size of families found this way is described in the Supplement. For the largest families in the UK Biobank, we display the intra-family PGS distribution alongside the population PGS distribution in **Figure 5**.

**Figure 5:**
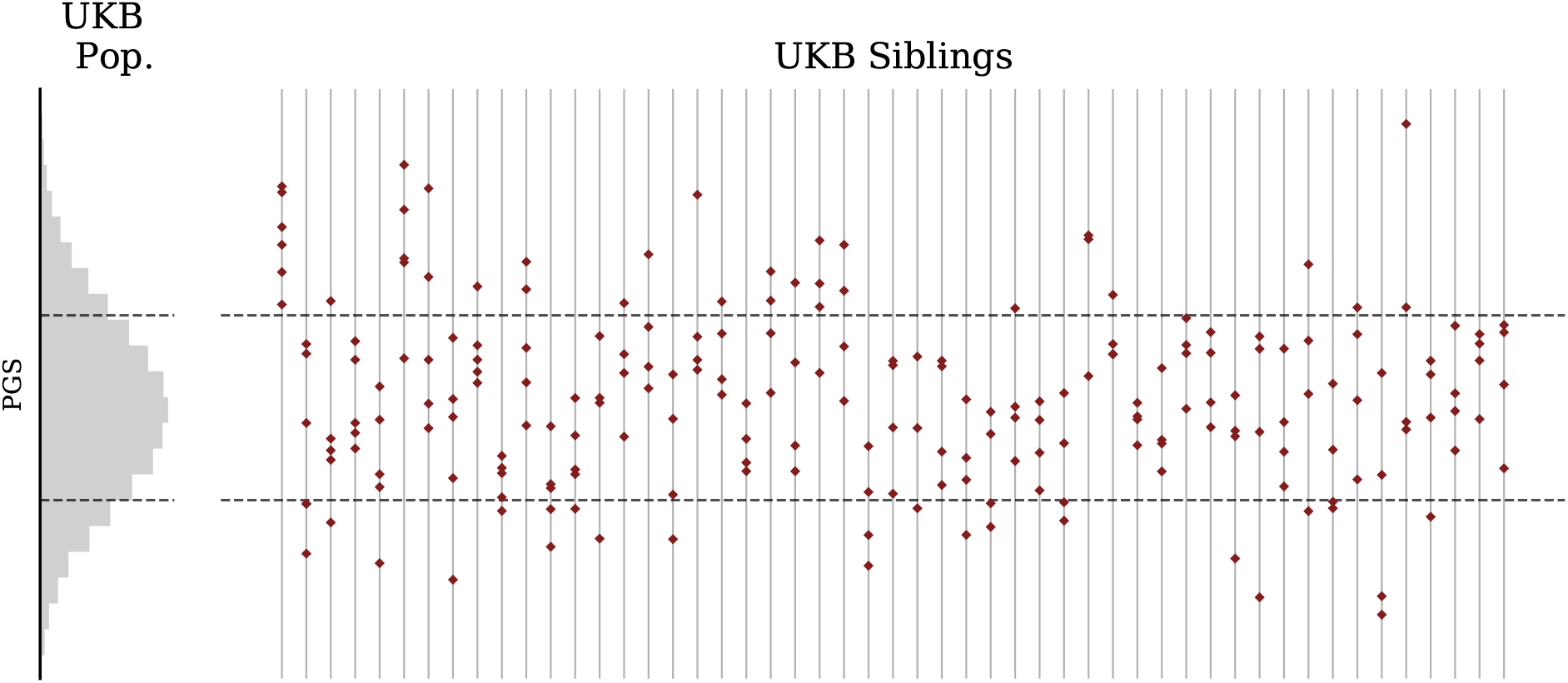
Type 2 Diabetes PGS by family (uncentered) illustrates significant variation within each family. Histogram on left describes UKB general population. Sets of siblings organized by family are displayed on the right to indicate both interand intra-family variation as compared to the general population. Note in some families all siblings are above or below the general population average in PGS. Horizontal lines indicate the standard deviation of the general UKB population.

Figure 3. and **Figure 5** show that some of the variation in the population PGS variance is due to inter-family variation and that the intra-family variation is reduced in comparison. For illustration we use the Type 2 Diabetes polygenic risk score. In **Figure 6**, we center PGS scores by family around the average PGS of its members. For the largest families, we display the population distribution alongside the centered PGS distribution and the re-centered distribution for each family. This demonstrates that, within a particular family, difference in intra-family polygenic score has close to 1/2 the variance of the general population which includes both an intra- and inter-family variation.

**Figure 6:**
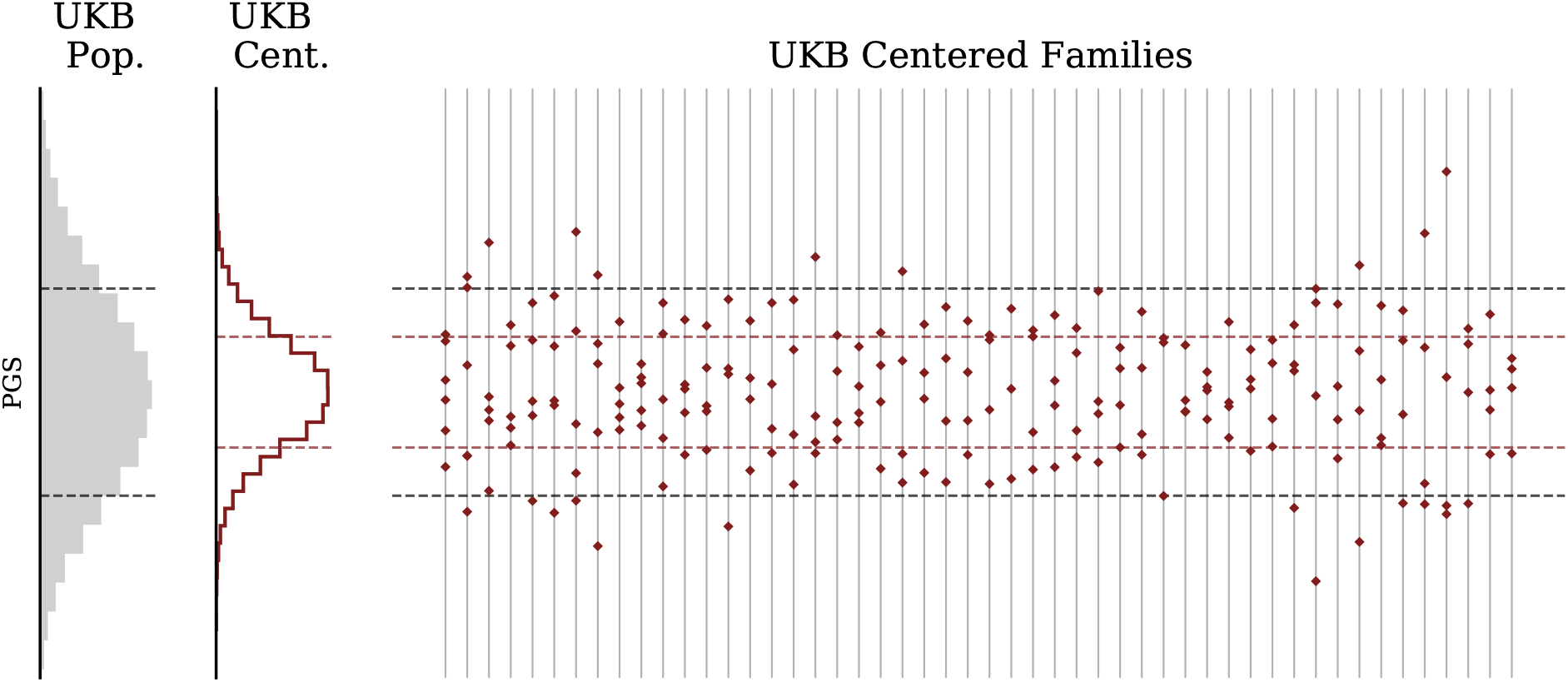
Type 2 Diabetes Polygenic Risk Score for UKB families (centered). Histogram on far left describes UKB general population. Middle histogram describes sibling scores after recentering relative to sibling average score (parental midpoint is generally not available). Sets of siblings organized by family are displayed on the right to indicate intra-family variation as compared to the general population. The sibling values have been centered within each family, unlike in the previous figure.

**Table 2:** Descriptive statistics for disease condition PGS: The ratio of unrelated (*U*) vs sibling (*S*) standard deviations is seen be 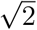 for essentially all disease PGS. *σ*(Δ*U*) is the standard deviation of the PGS-difference among unrelated individuals and *σ*(Δ*S*) is the corresponding standard deviation among siblings. PGS-corr. refers to correlation in polygenic score between siblings, and *m* is the implied assortativity. The analysis was performed on *N* = 21, 709 pairs of siblings/unrelated individuals. All uncertainties, listed in parentheses, are standard errors. *Note the population distributions for Gout and T1D deviate substantially from Gaussian (both have very sparse genetic architecture), so the derived values in the table should be interpreted with caution.

| Phenotype | PGS-corr. | $m$ | $\sqrt{2} \frac{\sigma(\Delta S)}{\sigma(\Delta U)}$ |
| --- | --- | --- | --- |
| Alzheimer's | 0.498<sub>(0.006)</sub> | 0.00<sub>(0.02)</sub> | 1.012<sub>(.007)</sub> |
| Asthma | 0.440<sub>(0.006)</sub> | -0.12<sub>(0.01)</sub> | 1.056<sub>(.007)</sub> |
| Atrial Fibrillation | 0.454<sub>(0.006)</sub> | -0.09<sub>(0.01)</sub> | 1.041<sub>(.007)</sub> |
| CAD | 0.454<sub>(0.006)</sub> | -0.09<sub>(0.01)</sub> | 1.044<sub>(.007)</sub> |
| Gout* | 0.390<sub>(0.006)</sub> | -0.22<sub>(0.01)</sub> | 1.108<sub>(.008)</sub> |
| Heart Attack | 0.482<sub>(0.006)</sub> | -0.04<sub>(0.02)</sub> | 1.019<sub>(.007)</sub> |
| High Cholesterol | 0.432<sub>(0.006)</sub> | -0.14<sub>(0.01)</sub> | 1.064<sub>(.007)</sub> |
| Hypertension | 0.435<sub>(0.006)</sub> | -0.13<sub>(0.01)</sub> | 1.066<sub>(.007)</sub> |
| IBD | 0.506<sub>(0.006)</sub> | 0.01<sub>(0.02)</sub> | 0.999<sub>(.007)</sub> |
| Ischemic Stroke | 0.509<sub>(0.006)</sub> | 0.02<sub>(0.02)</sub> | 0.990<sub>(.007)</sub> |
| Obesity | 0.484<sub>(0.006)</sub> | -0.03<sub>(0.02)</sub> | 1.025<sub>(.007)</sub> |
| Schizophrenia | 0.530<sub>(0.006)</sub> | 0.06<sub>(0.02)</sub> | 0.965<sub>(.007)</sub> |
| Type 1 Diabetes* | 0.292<sub>(0.006)</sub> | -0.42<sub>(0.01)</sub> | 1.189<sub>(.008)</sub> |
| Type 2 Diabetes | 0.444<sub>(0.006)</sub> | -0.11<sub>(0.01)</sub> | 1.049<sub>(.007)</sub> |

### 2.3 PGS results in Embryo Families

The distribution of PGS for all embryos (including different families) is similar to the PGS distribution to the UKB general population. Within a specific IVF family, the embryo PGS distribution is centered around the midparent PGS. In **Figure 7**, we display the general UK Biobank PGS distribution, the GP parent-embryo distribution, and the scores of individuals (organized by family) which make up the parent-embryo cohort.

**Figure 7:**
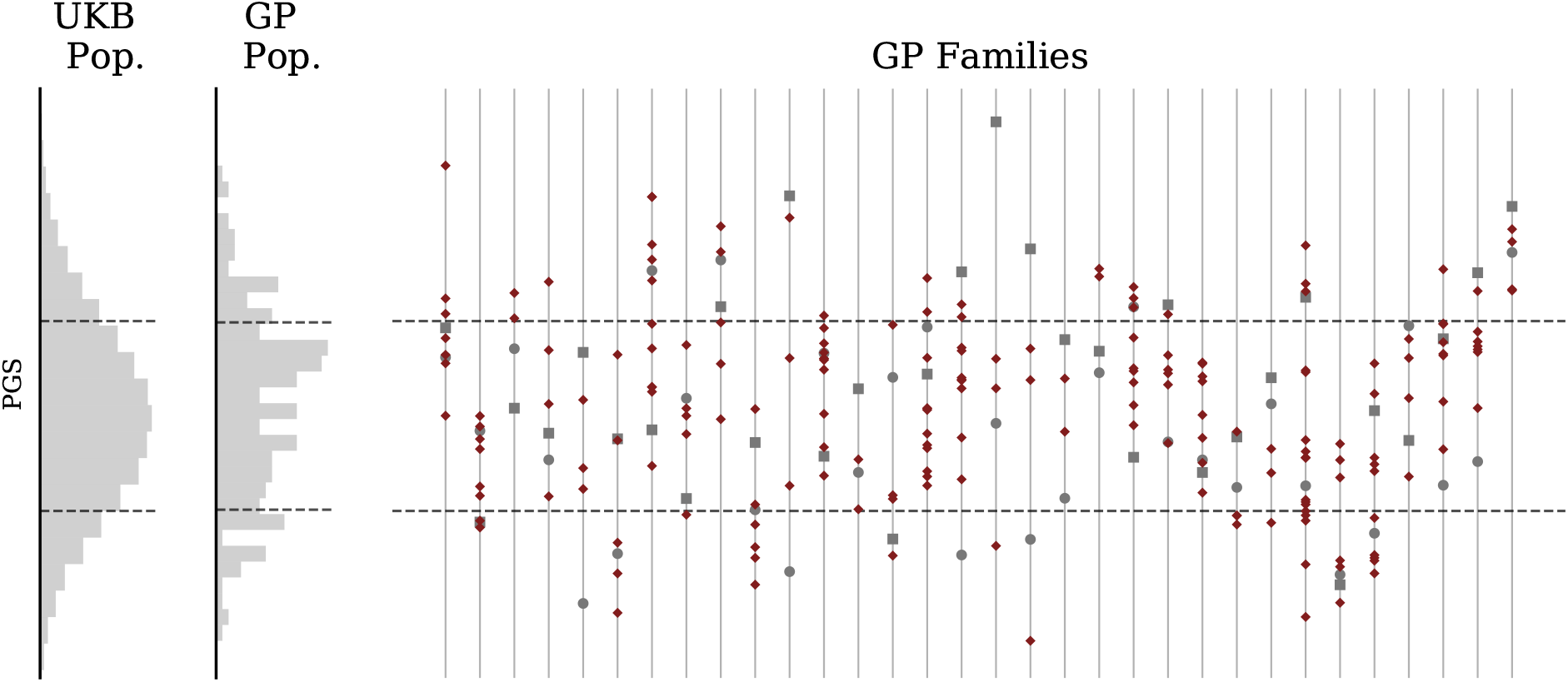
Type 2 Diabetes Polygenic Risk Score for GP/IVF families (uncentered). Far left histogram describes UKB general population. Middle histogram describes GP IVF embryos in aggregate (note this is a mixture of highly-related (sibs) (sibs from different families). Individual families are displayed on the right with mother represented by a circle and father by a square. Note siblings can have higher or lower PGS than either parent. Horizontal lines indicate standard deviations in the general populations.

As before, the variation in the cohort has an inter- and intra-family component. However, for this cohort we have access to the PGS of the parents for each embryo. We next demonstrate that distribution of embryo PGS is distributed around the midparent value which leads to the inter-family variation. In **Figure 8**, we display the general parent-embryo distribution alongside the midparent-centered parent-embryo distribution, along with the centered family in the parent-embryo cohort. Note that even when parental PGS differences are small, the variation within a set of embryos remains substantial.

**Figure 8:**
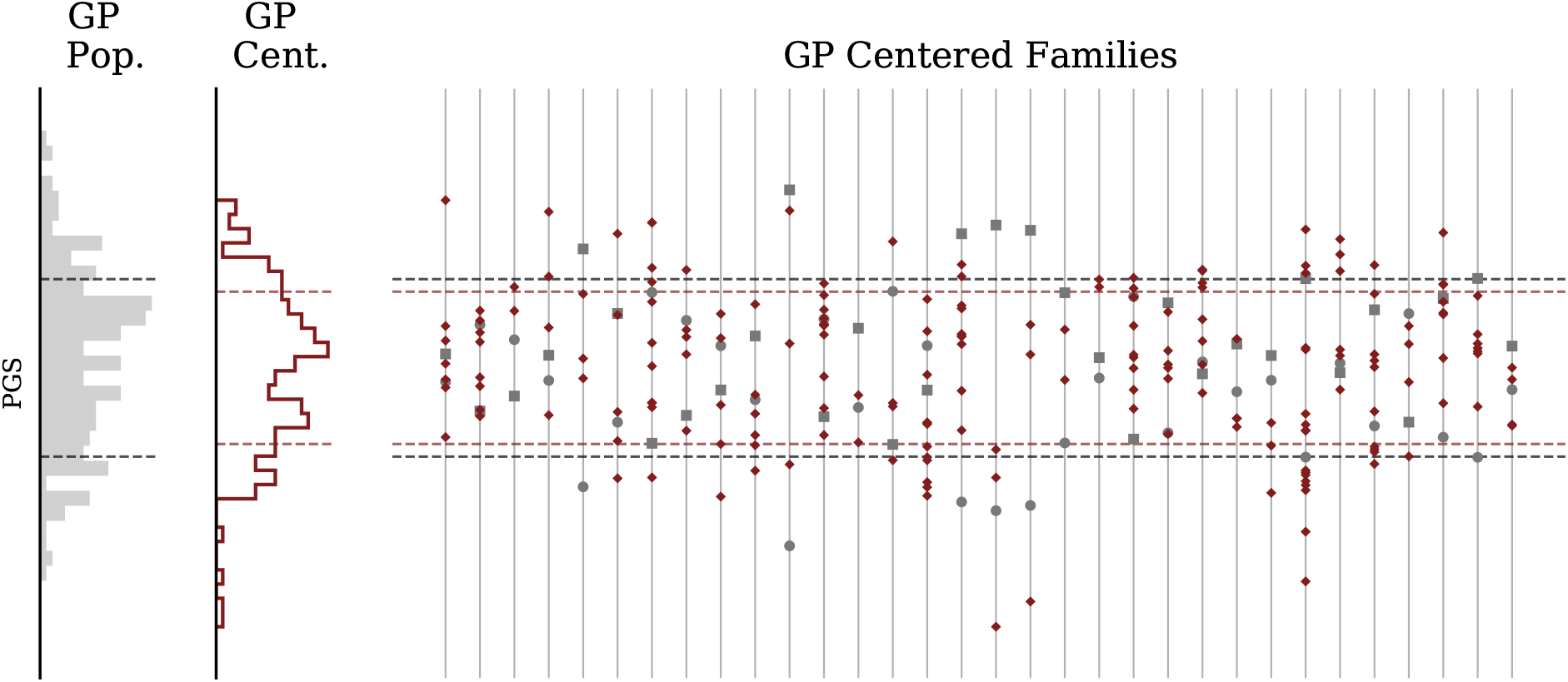
Type 2 Diabetes Polygenic Risk Score for GP/IVF families (centered). Far left histogram describes GP embryos in aggregate. Middle histogram describes GP IVF embryos after recentering with respect to parental midpoint. Individual families are displayed on the right with mother represented by a circle and father by a square. Families on right are displayed after recentering at midparent PGS. Again, the population standard deviations are marked with horizontal lines.

Figure 9. shows the Genetic Health Index score (as described in [43]) distributions in the UK Biobank compared to GP parental (saliva) and embryo samples. **Figure 10** and **Figure 11** use the Health Index in place of the Type 2 Diabetes polygenic score, displaying distributions within the UK Biobank and the parent-embryo cohort, as well as individual scores organized by fam ily. Health Index is a nonlinear function of genotype (as we discuss further below) so the 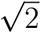 relationship between general population and within-family standard deviation does not necessarily hold.

**Figure 9:**
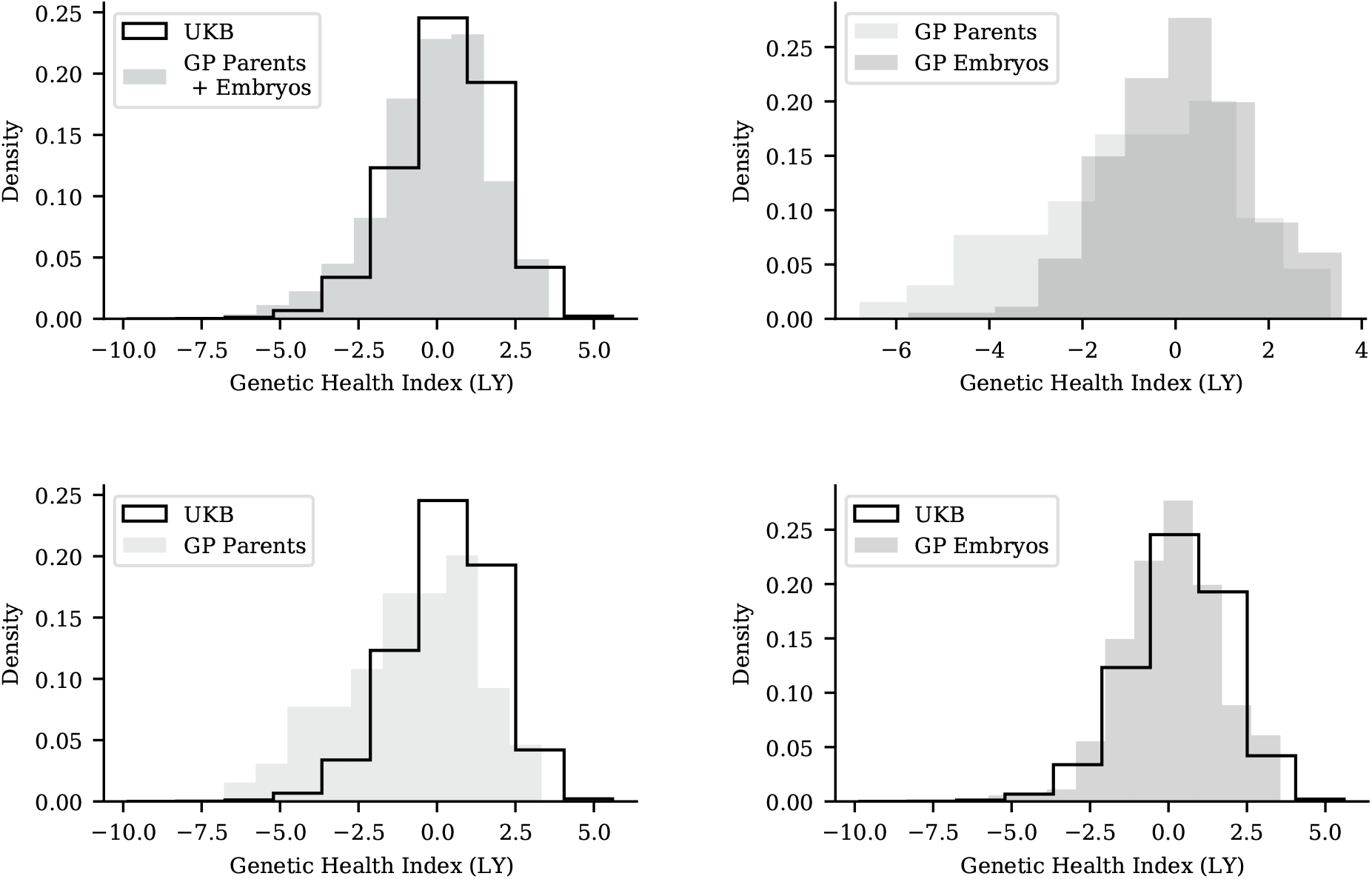
Genetic Health Index value distributions in the UK Biobank are similar to those of GP parents and embryos. Top-left: UK Biobank vs all GP samples. Top-right: GP parent samples vs GP embryo samples. Bottom-left UK Biobank vs parent samples. Bottom-right: UK Biobank vs embryo samples.

**Figure 10:**
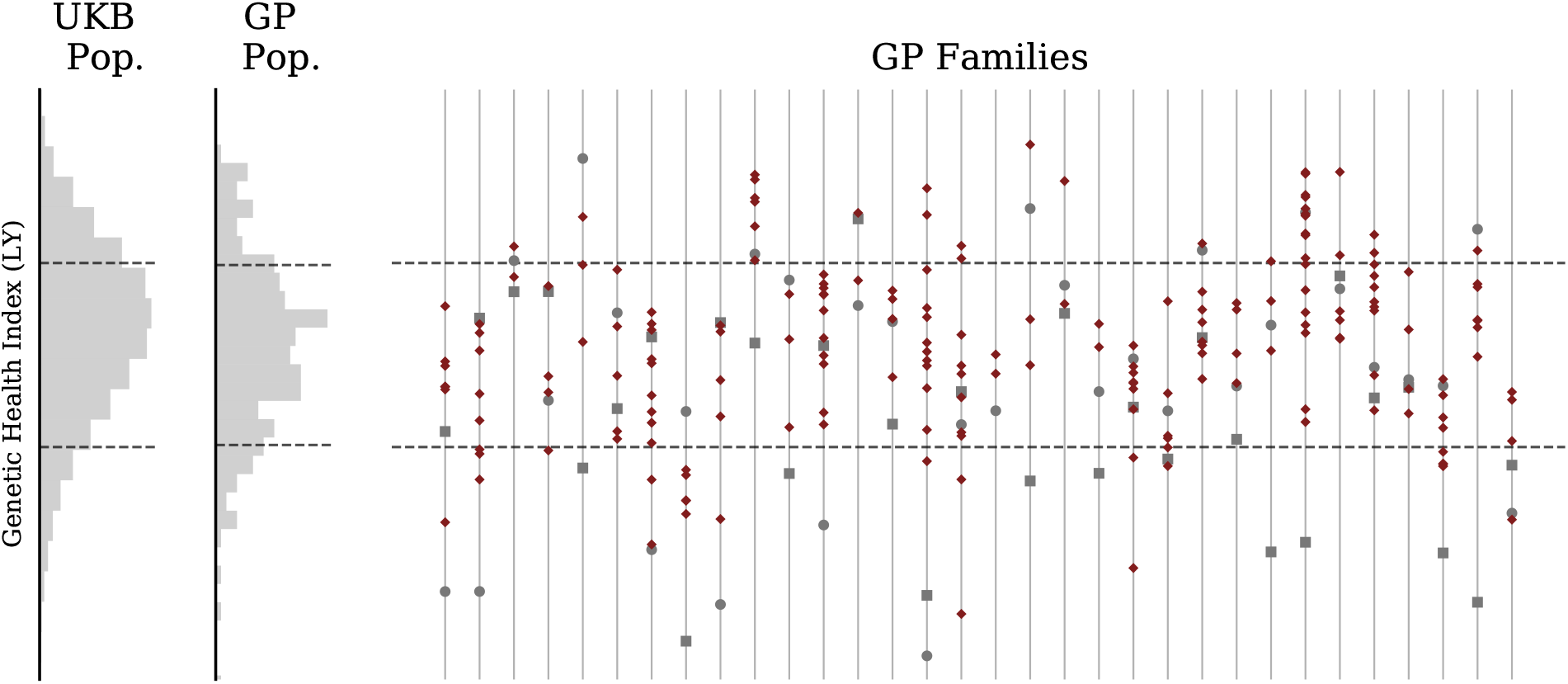
Genetic Health Index for GP/IVF families (uncentered). Far left histogram describes UKB general population. Middle histogram describes GP IVF embryos in aggregate (note this is a mixture of highly-related (sibs) (sibs from different families). Individual families are displayed on the right with mother represented by a circle and father by a square. Note siblings can have higher or lower Index than either parent

**Figure 11:**
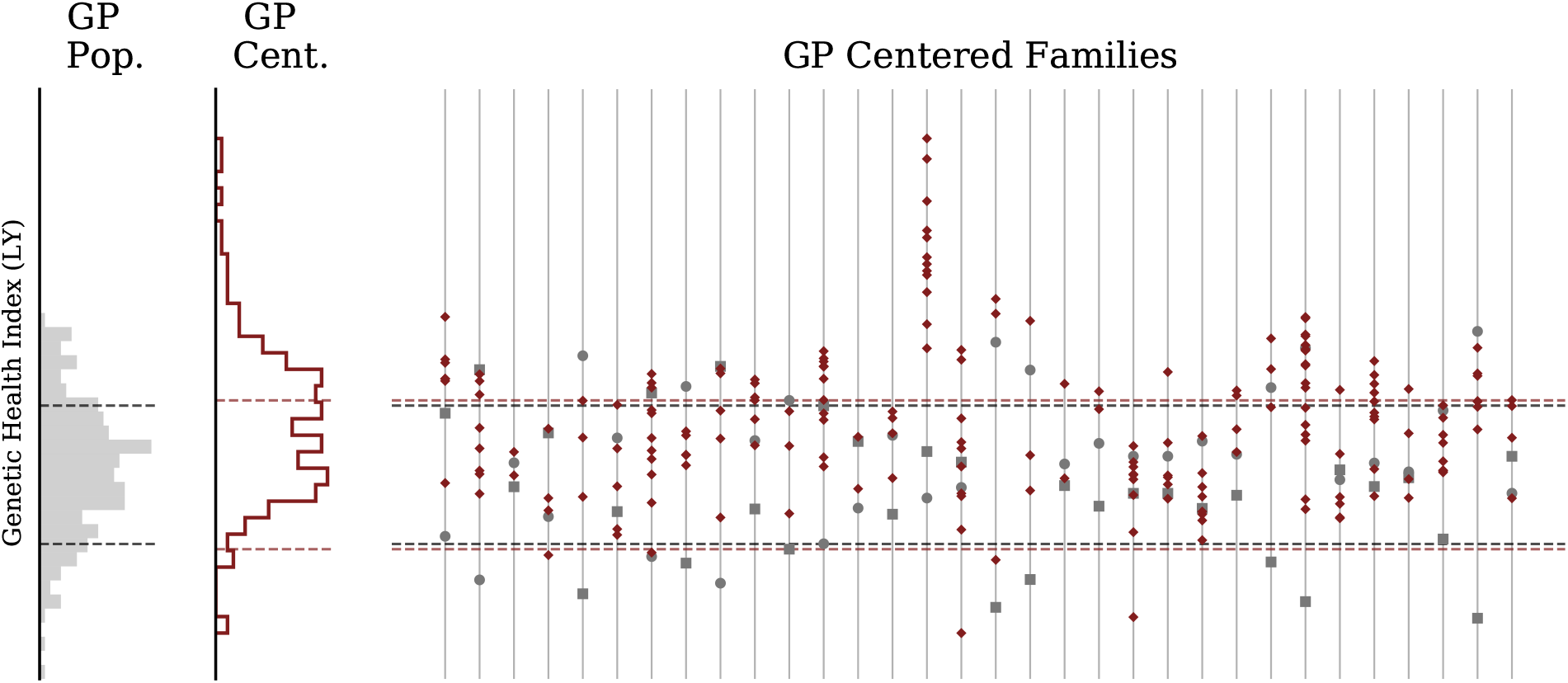
Genetic Health Index for GP/IVF families (centered). Far left histogram describes GP embryos in aggregate. Middle histogram describes GP IVF embryos after recentering with respect to parental midpoint. Individual families are displayed on the right with mother represented by a circle and father by a square. Families on right are displayed after recentering at midparent Index.

Among the families displayed in these figures, at position number 15 from the left, we encounter an interesting case of sibling polygenic distribution relative to the parents. In the family all siblings have significantly higher Health Index score than the parents. This arises in an interesting manner: the mother is a high-risk outlier for condition X and the father is a high-risk outlier for condition Y. (We do not specify X and Y, out of an abundance of caution for privacy, although the patients have consented that such information could be shared.) Their lower overall Health Index scores result from high risk of conditions X (mother) and Y (father). However, the embryos, each resulting from unique recombination of parental genotypes, are normal risk for both X and Y and each embryo has much higher Health Index score than the parents. This can be discerned from detailed examination of individual genomes and associated polygenic scores.

It is worth noting that the Health Index sums absolute risk (probability of getting the condition) per condition times lifespan impact. Absolute risk is a nonlinear function of polygenic score, whereas all polygenic scores used in this paper (and almost all in the published literature) are linear in the genotype. In this case the absolute risks of the parents for X (mother) and Y (father) are very high. Embryo genomes, due to the diploid nature of human sexual reproduction, are a blend of maternal and paternal genotypes. In this case all of the polygenic scores of the embryos cluster around the parental midpoint values for both X and Y. Due to nonlinearity, the absolute risks for X and Y of the embryos are all much lower than for the parents, and the embryos have higher Health Index scores than the parents.

### 2.4 Detection of Recombination Events

Results are obtained using the method described in section 4.3, which identifies recombination events using statistical properties of paired sibling genotypes. We apply the algorithm of section 4.3 to imputed UKB data. In **Figure 12**, we display the cumulative number of breaks from the sibling algorithm compared to the results from deCODE [46] for chromosomes 1 and 22. **Figure 13** are histograms of breaks by location on chromosomes 1 and 22. Known hot spots [46] (regions with very high recombination activity) are readily visible. These are average results from 21k self-identified European sibling pairs. Similar results for the remaining autosomal chromosomes are presented in the Supplement.

**Figure 12:**
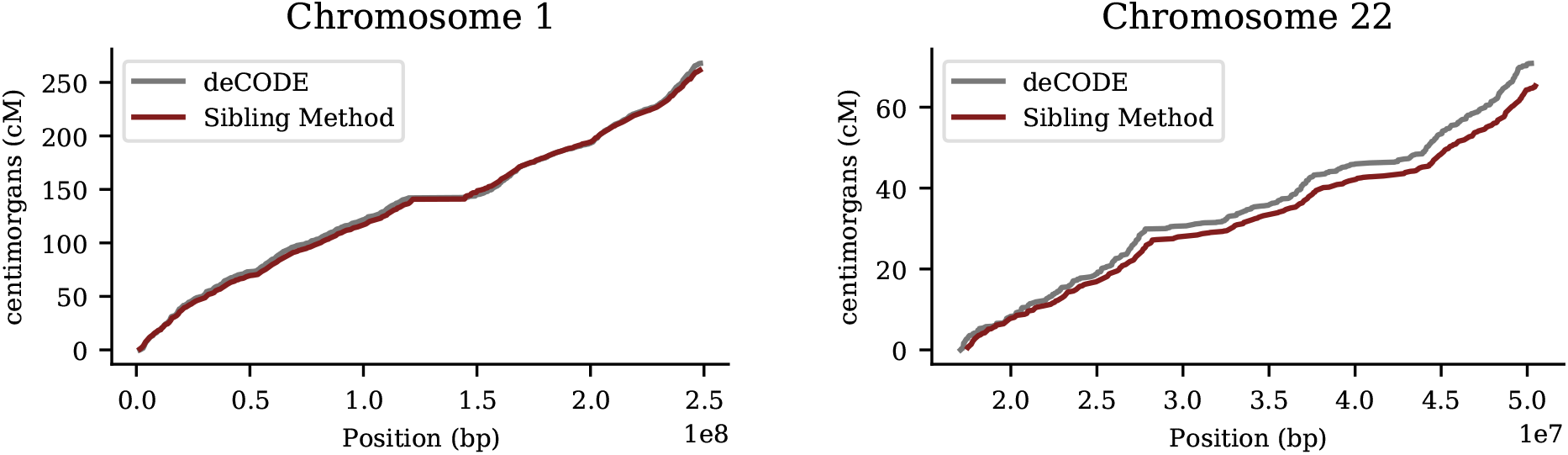
Cumulative recombination length of chromosomes 1 and 22. (i.e., total Morgan length). Red line is the siblings-pair method applied to 21k European ancestry sibs from UK Biobank. Gray line is from deCODE (Science 2019) result using population of Iceland.

**Figure 13:**
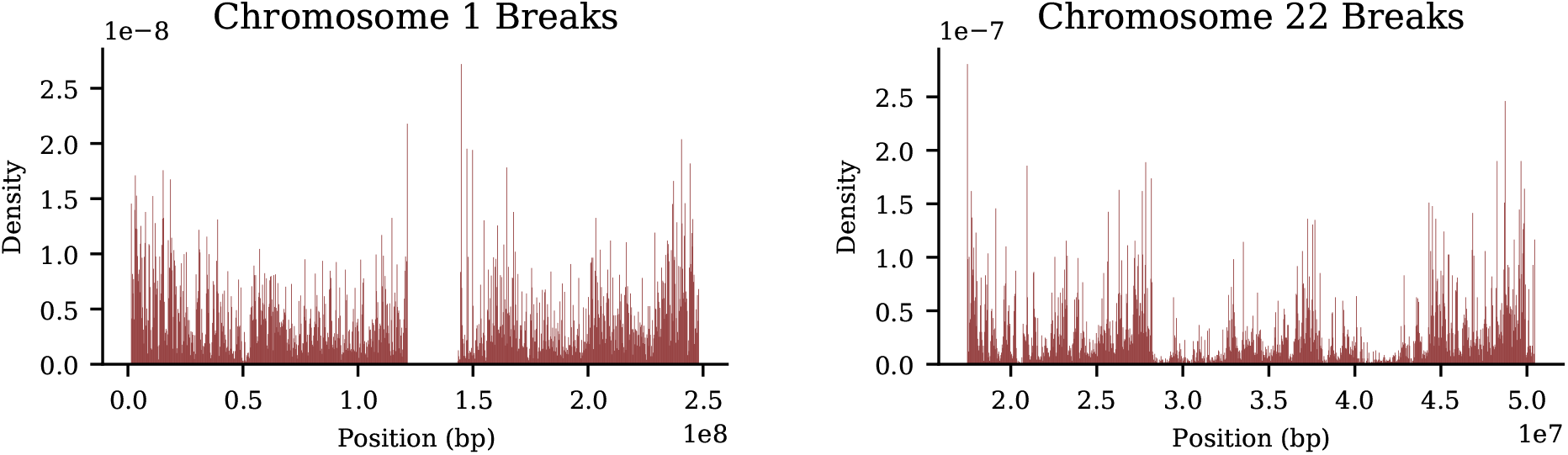
Density of recombination breaks on chromosomes 1 and 22 from sibling-pair method, computed using 21k European sibling pairs.

Agreement between sibling-pair method and deCODE results is very strong. In particular, the features in plots of cumulative breaks per chromosome are clearly similar in both sets of results. In the Supplement we discuss three anomalies (specific regions of high recombination activity in the sibling-pair results, relative to deCODE) found on chromosomes 2, 15, and 21 that warrant further investigation in the future.

Because our method uses flanking windows for statistical comparisons, breaks which are very close to the ends of chromosomes (i.e., telomere regions, where recombination rates are generally high) will be under counted. This observation also affects methods which track the state of parental hetero- and homozygous sites in offspring. It is difficult to know which methods more severely over or under count breaks in the endpoint regions.

It is important to note that even slightly inaccurate maps of recombination breaks are still useful for machine learning methods which attempt to predict local recombination rate based on DNA motifs in the region. From the machine learning perspective, the impact of modest inaccuracies is simply a small decrease in statistical signal. The over-representation of certain motifs or specific mutations in hotspot regions should still be detectable by automated learning methods. The primary benefit from the sibling-pair method is a very large expansion in the size (and ancestry group coverage) of available maps (catalogs) of recombination events across the genome.

In **Table 3**, we summarize cumulative results for each chromosome generated from the sibling method and from deCODE. Note that deCODE provides the cM length as the average over parents – which results in 1/2 the number of expected breaks – while the sibling-pair method is designed to detect 1/2 of all breaks (i.e., the boundaries between type 1 and type 2 regions; see section 4).

**Table 3:** Recombination length (centiMorgans) by chromosome. obtained from 21, 514 UKB European ancestry sibling pairs, compared to deCODE (Iceland) results. Standard error for sibling-pair method given in parenthesis.

| Chr. | Sib. method | deCODE | Chr. | Sib. method | deCODE |
| --- | --- | --- | --- | --- | --- |
| 1 | 261.4<sub>(0.6)</sub> | 267.8 | 12 | 159.7<sub>(0.5)</sub> | 165.5 |
| 2 | 266.4<sub>(0.7)</sub> | 251.8 | 13 | 122.7<sub>(0.4)</sub> | 127.2 |
| 3 | 212.4<sub>(0.6)</sub> | 218.3 | 14 | 114.1<sub>(0.4)</sub> | 116.0 |
| 4 | 202.1<sub>(0.6)</sub> | 202.9 | 15 | 121.5<sub>(0.4)</sub> | 117.3 |
| 5 | 194.5<sub>(0.6)</sub> | 197.1 | 16 | 121.3<sub>(0.4)</sub> | 126.6 |
| 6 | 186.8<sub>(0.6)</sub> | 186.0 | 17 | 119.6<sub>(0.4)</sub> | 129.5 |
| 7 | 174.9<sub>(0.5)</sub> | 178.4 | 18 | 109.9<sub>(0.4)</sub> | 116.5 |
| 8 | 162.1<sub>(0.5)</sub> | 161.5 | 19 | 98.3<sub>(0.4)</sub> | 106.4 |
| 9 | 157.4<sub>(0.5)</sub> | 157.4 | 20 | 98.7<sub>(0.4)</sub> | 107.8 |
| 10 | 162.6<sub>(0.5)</sub> | 169.3 | 21 | 75.4<sub>(0.3)</sub> | 62.9 |
| 11 | 155.0<sub>(0.5)</sub> | 154.5 | 22 | 65.2<sub>(0.3)</sub> | 70.8 |
|  |  |  | Total: | 3,338.0<sub>(3.3)</sub> | 3,391.4 |

Although the UKB cohort is predominantly self-reported European, there are a non-trivial number of sibling pairs with other self-reported ancestry. The question of variation in recombination rate by self-reported ancestry group can be investigated, at least at a preliminary level, using the non-European siblings found in UKB. Application of the new method to UKB sibling pairs yields the following whole genome recombination lengths for the three largest (self-reported) ancestry groups: Europeans (21, 514 pairs) 3,338 cM, South Asian (292 pairs) 3,073 cM, African (174 pairs) 3,453 cM. If we adopt the European distribution as the null hypothesis – mean 33.38 breaks per individual in a pair, SD = 4.8 – we obtain the following results: 1. The South Asian deviation in average is 2.65 fewer breaks (with N = 292). In null model SD units this is 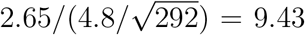, so statistically significant. 2. The African deviation in average is 1.15 more breaks (N = 174). In null model SD units this is 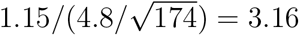, so statistically significant.

We plan to investigate this topic in the future with more non-European sibling data as it becomes available. It is possible that the differences in total recombination rates found above are the result of some kind of systematic error. For example, the sibling-pair method might exhibit higher or lower recombination detection rates due to differences in how genome imputation works across different ancestry groups.

## 3 Discussion

Roughly half of the genetic variation in a population is within-family, and the other half between families. This implies that the standard deviation of sibling polygenic score distributions is roughly 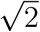 times smaller than for the general population. Siblings are more similar to each other than randomly selected individuals in the population, but there is still significant variation within each family. We have explored these, and related, consequences of simple population genetics using the power of modern genotyping, large biobanks, and a unique dataset of IVF families.

In earlier work it was shown that PGS perform almost as well in differentiating siblings by disease status or quantitative trait value as in pairs of unrelated individuals [27, 44]. It was also shown that expected gains from selection on PGS or Health Index score are substantial [43]. For the latter result to hold there must be significant variance among siblings on the relevant PGS. In this paper we exhibited this variation using many traits.

We have also developed a new technique that directly probes DNA recombination – the molecular mechanism responsible for sibling genetic differences. This new sibling-pair method can be applied to large datasets with many thousands of sibling pairs. In this project we created a map of roughly 1.44 million recombination events using UKB genomes. Similar maps can now be created using other biobank data, including in non-European ancestry groups that have not yet received sufficient attention. The landmark deCODE results were obtained under special circumstances: the researchers had access to data resulting from a nationwide project utilizing genealogical records (unusually prevalent in Iceland) and widespread sequencing. Using the sibling-pair method results of comparable accuracy can be obtained from existing datasets around the world – e.g., national biobanks in countries such as the USA, Estonia, China, Taiwan, Japan, etc.

It seems possible that deep learning approaches using this training data could identify new DNA motifs that are associated with increased or decreased local recombination probability. This may advance our molecular understanding of meiosis, a fundamental biological process. Regarding applications, we note that many chromosomal abnormalities are the result of errors introduced through recombination. Improvements in computational analysis of embryo genotypes may result in better accuracy in embryo screening against aneuploidy.

## Methods and Data

### UK Biobank and GP Biobank

The two datasets used in this work are the 2018 release of the UKB and a cohort of parent and embryo genotypes from families undergoing preimplatation genetic testing for polygenic (PGT-P) screening at GP.

The GP samples are processed on the same (Affymetrix) SNP array platform which is used by the UKB. The details of the genotyping procedure and quality control are described at length in [47]. We restrict all samples to those with European ancestry with a total of 32 individual parent-embryo groups. Pre-implantation genetic testing for aneuploidy was carried out on all embryos and only euploid embryos were considered for analysis in this work. GP data was collected along internal GP guidelines, conforming with local legislation and institutional requirements. The patients/participants provided their written informed consent to participate in this study. Written informed consent was obtained from the individual(s) for the use of any potentially identifiable data included in this article, although no identifiable information was used, nor made available to researchers.

UKB data was collected under policies conforming with national and requirements and subject to privacy rights. (https://www.ukbiobank.ac.uk/privacy-policy) All data handled by researchers in this work was de-identified.

MSU researchers working on this project do not have access to any identifiable data and are covered under MSU IRB [STUDY00006493].

For the UK Biobank, all of our samples are restricted to self-reported European (in UKB terminology, white) ancestry unless otherwise stated. The details of the genotyping and quality control steps are found in the supplements, section 1.

Polygenic predictors are obtained through training on either the UK Biobank or a wholly separate GWAS. The testing set we use is the set of 21k sibling pairs in the UK Biobank (*∼*40k individuals) – all of whom have been withheld from predictor training and tuning. The majority of predictors are trained using the LASSO algorithm in scikit-learn. The quantitative trait predictors are described in detail in [3, 27], while case-control predictors are described in [44]. Several predictors are trained using PRS-CS and publicly available GWAS which have a much higher case-count than available in the UK Biobank. Details of the training are described in the Supplement of [43] and we provide a brief summary in the Supplement.

### 4.2 Polygenic scores and classical results from population genetics

In this section we review some classical results of population genetics with an emphasis on how they relate to polygenic scores. A phenotype is typically modeled (assuming no *G × E* interactions) as a linear combination of genetic and environmental effects as

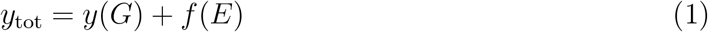

where *G, E* are the genetic and environmental components. The genetic contribution to a particular phenotype is known as the heritability and is defined as

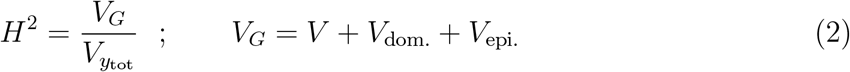

where *V* denotes the additive variance. The genomic variance is typically separated into additive, dominance and epistasis components. We are from now on focusing on the additive component only and denote it as *V*, without any subscripts. The linear component of heritability is referred to as the narrow-sense heritability and is given by

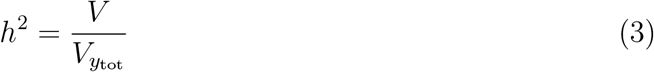

i.e., it is just the additive component of variance. In this work, we assume additive models such that the genetic contribution to the phenotype of an individual *i* is

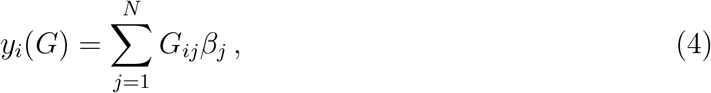

where *β*_*j*_ is the effect size for a given SNP *j*. With this definition we can estimate the narrow-sense heritability by a second method

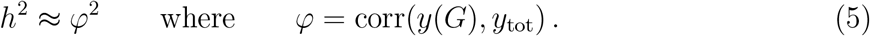

In **Table 1** the average of both methods is recorded.

The covariance between sibling polygenic scores *S*_1_ and *S*_2_ can be calculated, assuming additivity, as

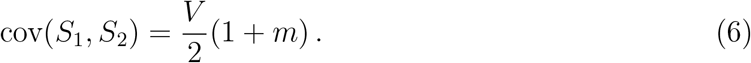

Here *m* is the assortivity, or the degree to which mates resemble each other on this specific polygenic score (*m* = 0 for random mating) [48].

For a given pair of siblings the sum over genetic loci can be separated into the subset which are IBD and another subset which vary randomly as in the general population (neglecting assortation):

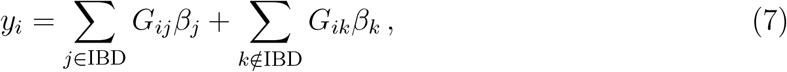

so that the difference in sibling values is

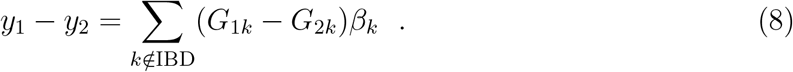

Since on average half the SNPs are IBD and that variance is additive, the variance in the difference between siblings Δ*S*, which arises from the loci that are *not* IBD will be half that found for the difference between unrelated individuals Δ*U* :

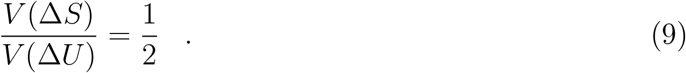

Again, we have neglected assortation. Slight deviations from this result are observed for some traits for which assortation is significant. Consequently the standard deviation in the distribution of sibling differences will be 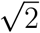 times smaller than for unrelated pairs of individuals. Similarly, one can show that the distribution of sibling P GS values, which is centered at the parental midpoint, has standard deviation which is 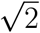 times smaller than the distribution of PGS values in the general population [49].

Note in **Figure 5** and **Figure 6** (based on UKB data) we do not have access to parental midpoint values and hence plot the sibling distribution relative to the average among the available sibs. This is subtly different from recentering using parental midpoint (the average among sibs only approaches parental midpoint when number of sibs is large, and in the UK data this is usually not the case), and further reduces the variance of the resulting distribution. The 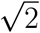 effect (reduction in variance among sibs) from population genetics is easier to evaluate using *differences* in sibling scores, because parental midpoint is not required.

#### 4.2.1 Binary traits and the Genetic Health Index

The discussion until this point has assumed quantitative phenotypes. For case-control phenotypes, several approaches exist in the literature. For the context of this work, the mixed-distribution model of polygenic scores is adopted where both cases/controls follow normal distributions but are shifted from one-another. This is written as

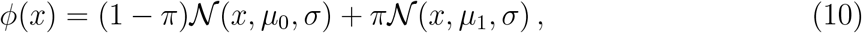

where *x* is the PGS, *µ*_0*/*1_ is the mean PGS of controls/cases, *σ* is the standard deviation of PGS among the controls (The standard deviations for cases and controls do not have to be equal in principle but are usually very close in practice. The model assumes them equal and we use *σ*_0_ due to better statistics.) and *π* is the lifetime risk. The mixed-distribution model can be translated into risk as follows

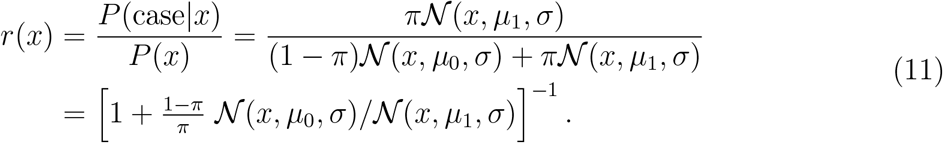

With this model of risk, we can construct a quantity which describes expected lifespan gain for an individual *i* given by

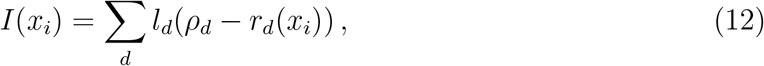

where *ρ*_*d*_ is the average lifetime risk of a particular disease *d* and *l*_*d*_ is the average lifespan reduction of that disease in units measured in years (life-years, LY). This quantity is referred to as a Genetic Health Index. Clearly this is only a proxy for lifespan gain as it does not account for disease covariances and cannot possibly include lifespan impacts from all phenotypes. For further discussion about properties and behavior of this Genetic Health Index, see [43].

### 4.3 Detection of Recombination Events Using Sibling Pairs

Here we describe a novel algorithm for detecting recombination events using sibling pairs. This sibling-pair method does not require parental genotypes. Let *s*_1_ and *s*_2_ be the two sibling genotypes. Consider a specific SNP location and define two DNA windows of size *N* flanking this locus (**Figure 14**). (We may refer to one window *w*_*−*_ on the “left” of the reference SNP and the other *w*_+_ to the “right”.) The algorithm computes the similarity of genotypes *s*_1_ and *s*_2_ in each of the flanking windows. For example, the similarity could be defined as simply the number *N*_*±*_ of loci in each window which are identical in state in both siblings (*N*_*±*_ are each numbers between 0 and *N*). We can then define an indicator function *Z*(*N*_+_, *N*_*−*_) (see below) which is used to detect discontinuities in similarity, comparing *w*_+_ with *w*_*−*_.

**Figure 14:**
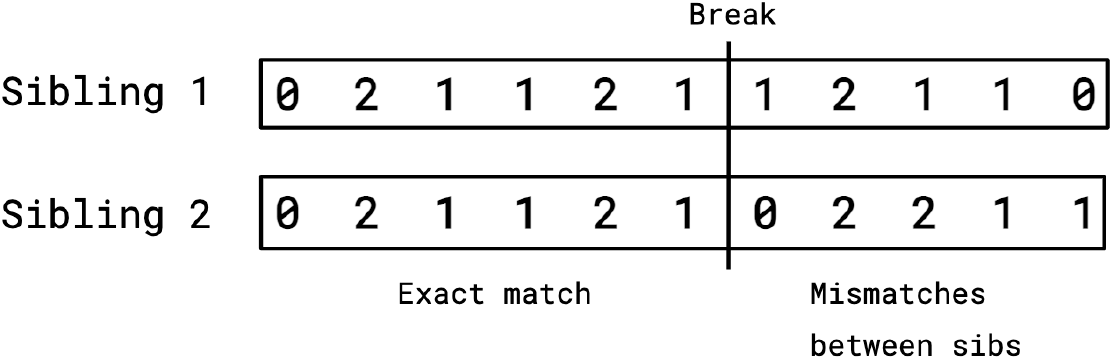
The recombination algorithm detects breaks between regions in which both siblings have identical SNPs and regions where they differ. SNP genotypes for sibling 1 and 2 are shown as counts of the major alleles. The recombination break occurs at the boundary between regions. On the left both siblings have inherited the same chromatid strands from the parents. A recombination break, which could occur in either the egg or the sperm progenitor of either sibling, causes mismatches in the SNP genotypes in the region on the right – i.e., on the right the siblings no longer have the same chromatid strands.

For a given window, there are 3 distinct types of region for a sibling pair. Let the father’s chromosome strands (chromatids) be *Y*_*a*_, *Y*_*b*_ and the mother’s chromosome strands be *X*_*a*_, *X*_*b*_, and adopt the notation *s*_1_ : *s*_2_ to describe the genotype of each of the siblings at the same position. Then, the state of a specific region in the sibling pair must realize one of the following possibilities:

Identical (type 1): *X*_*a*_*Y*_*a*_ : *X*_*a*_*Y*_*a*_, *X*_*b*_*Y*_*b*_ : *X*_*b*_*Y*_*b*_, *X*_*a*_*Y*_*b*_ : *X*_*a*_*Y*_*b*_, *X*_*b*_*Y*_*a*_ : *X*_*b*_*Y*_*a*_ (4 possibilities)

Partially identical (type 2): *X*_*a*_*Y*_*a*_ : *X*_*a*_*Y*_*b*_, *X*_*b*_*Y*_*b*_ : *X*_*a*_*Y*_*b*_, *…* (8 possibilities)

No IBD segments (type 3): *X*_*a*_*Y*_*a*_ : *X*_*b*_*Y*_*b*_, *X*_*b*_*Y*_*b*_ : *X*_*a*_*Y*_*a*_, *X*_*a*_*Y*_*b*_ : *X*_*b*_*Y*_*a*_, *X*_*b*_*Y*_*a*_ : *X*_*a*_*Y*_*b*_ (4 possibilities)

From these observations, roughly one quarter of a sibling’s genome will match identically with the other. A recombination event can then be detected by scanning along the chromosome for regions in which both siblings agree completely (modulo rare genotyping errors) in *w*_*−*_ but differ from identical in *w*_+_. The location at which the identical (type 1 region) to non-identical (type 2 region) change happens is a recombination event.

Almost all breaks will be transitions from region type 1 to type 2 or region type 2 to type 3 because a jump from type 1 to type 3 would require a coincidence in meiosis between both the sperm and egg, which is highly unlikely. The sibling-pair method is designed to detect breaks from type 1 to 2 (or, in principle, 1 to 3) regions, but will not record changes from type 2 to 3. Therefore the sibling-pair method will detect 1/2 the total number of breaks.

We define an indicator function *Z* which signals discontinuities in statistical properties of flanking windows:

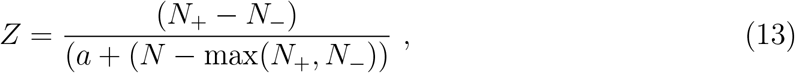

where *a* is a positive number much less than *N*, which keeps the denominator from ever becoming zero. This function has useful properties:

1. The numerator is large in magnitude when the *s*_1_ : *s*_2_ similarities in the two flanking windows are very different (e.g., if one window is in a type 1 region and the other is not).
2. The denominator is small when at least one of the flanking windows has close to maximal similarity *N*, which tends to occur in type 1 regions.

*Z* reaches a local maximum near a recombination break point, with *Z >* 0 indicating a transition from type 1 to 2 region (from “left” to “right”), while *Z* < 0 indicates a transition from type 2 to type 1 region.

The parameters *N, a* are tuned, and performance evaluated, using family data (both IVF and from UKB) in which parental genotypes are available. The parameters are chosen so that *Z* is robust against genotyping errors or mutations (which reduce similarity of IBD regions from 1), and clusters of low MAF SNPs (which can cause non-IBD regions to appear IBD). With parental genotypes we can make use of hetero- and homozygous loci to directly track which segments of chromatid have been passed to each sibling.

What is novel here is that recombination breaks can be detected using sibling pair data alone, once the indicator function *Z* has been properly adjusted. Many biobanks, including UKB, have large numbers of siblings in their cohort, whereas trios containing mother, father, and child are comparatively rare. Thus, this new technique will add significantly to the amount of data available for study of recombination. In this work we create a dataset of 1.44 million recombination events found in 21k UKB sibling pairs. We anticipate that datasets like ours will be useful in machine learning to discover the molecular mechanisms which control the local rate of recombination in the genome. For example, deep learning could be used to detect specific DNA motifs which are associated with recombination hotspots or regions where recombination is rare.

Note we assume that each recombination event can be considered as a draw from a probability distribution. Effects such as crossover interference (which are consequences of the underlying molecular mechanisms) cause breaks to be less likely to occur in close proximity to each other, violating the assumption that each individual break is an independent event. However, this is a small correction as with only 70 breaks across 22 chromosomes it is unlikely that two breaks occur close together. Since crossover interference does not inhibit a sib1 break conditional on a nearby sib2 break, the results from the sib-pair method detects very slightly more breaks in close proximity than occur in a single meiosis. However, as mentioned this is a very small effect as nearby breaks are rare.

## Data Availability

All data produced in the present study are available upon reasonable request to the authors

## 5 Acknowledgements

Computational resources provided by the Michigan State University High-Performance Computing Center. The authors acknowledge acquisition of data sets via UK Biobank Main Application 15326. The authors thank Jia Xu and Steve Hsu for discussions. Steve Hsu reviewed the manuscript and provided useful edits.

## 6 Data Availability

Access to the UK Biobank resource is available via application (http://www.ukbiobank.ac.uk). Genomic Prediction (GP) datasets generated for this study will not be made publicly available, in accordance with UKB and GP policy.

UKB data was collected under policies conforming with national and local requirements and subject to privacy rights. (https://www.ukbiobank.ac.uk/privacy-policy) All data handled by researchers in this work was de-identified.

GP data was collected along internal GP guidelines, conforming with local legislation and institutional requirements. The patients/participants provided their written informed consent to participate in this study. Written informed consent was obtained from the individual(s) for the use of any potentially identifiable data included in this article, although no identifiable information was used, nor made available to researchers. GP data is not available to researchers outside the company.

The 1.44 million recombination events found using the UKB siblings are available for use by other researchers www.github.com/MSU-Hsu-Lab/sibling-variation-paper-2022.

## 7 Author contributions

Authors, listed alphabetically according to last name, contributed in the following categories: Conceptualization, LL, TGR, EW; Methodology, MH, LL, TGR, EW; Software, MH, LL, EW; Validation, MH, LL, TGR, EW; Formal analysis, MH, LL, TGR, EW; Investigation, MH, LL, TGR, EW; Resources, LL, TGR, EW; Data curation, LL, EW; Writing—original draft preparation, LL, MH; Writing—review and editing, LL, TGR, EW; Visualization, LL, EW; Supervision, LL, EW; Project administration, LL; Funding acquisition, LL, TGR, EW; All authors have read and agreed to the published version of the manuscript.

## 8 Competing Interests

The authors declare the following competing interests: EW and LL are employees and shareholders of Genomic Prediction, Inc. MH is a volunteer (uncompensated) research intern at GP. TGR and MH declare no competing interests.

## Notes

### Funding Statement

This study did not receive any funding

### Author Declarations

Ethics IRB of Michigan State University waived ethical approval for this work. It was classified as non-human subjects because we only analyzed genomes with no identifying information.

### Summary of Updates

IVF family graphs have been modified slightly -- some aneuploid embryos were included in the earlier version graphs that should have been removed. Minor edits and clarification concerning the nonlinear nature of the Health Index and interpretation of family 15 scores.

