## supplementary material for "Sibling Variation in Phenotype and Genotype: Polygenic Trait Distributions and DNA Recombination Mapping with UK Biobank and IVF Family Data"

September 26, 2022

### 1 UK Biobank

#### 1.1 Sibling Set Identification

This section summarizes how sibling family structure was obtained. The UK Biobank provides pair relation data via Kinship coefficients and IBS0. Sibling pairs are identified by filtering for kinship coefficients >0.176 and IBS0 > 0.0012. This results in a set of 22,667 full sibling pairs, in agreement with [1], which identifies the number of sets of full trios and quartets (with both parents) but does not identify the number of sibling sets of larger than 2 (without parents).

Larger family structures exist in UK Biobank as pointed out in [1] and the relationship provided from the UK Biobank is in the form of pairwise data. From the set of full siblings, larger sets of (sibling-only) families are identified by collecting all sets such that if A-B and B-C are full sibling pairs, the superset A-B-C must all be full-siblings. Using this method, we identify 19,017 sets of 2; 1,051 sets of 3; 66 sets of 4; 11 sets of 5; and 2 sets of 6. Restricting to sets with only European self-reported status results in 18,239 sets of 2; 987 sets of 3; 61 sets of 4; 11 sets of 5; and 2 sets of 6.

Among the sibling pairs, restriction to different self-reported ethnicities results in 21,514 European pairs, 292 South Asian pairs, 174 African pairs and 30 East Asian pairs. In this work, we only consider non-European sibling pairs for the sibling-pair method results.

#### 1.2 Genotype Quality Control

In this work we use genotypes provided from the 2018 release of the UK Biobank which are genotyped on the Affymetrix UK Biobank Axiom and UK BiLEVE Axiom arrays [1]. The UK Biobank provides both directly called genotypes and a set of imputed genotypes. For the called genotypes we filter out SNPs with MAF <0.1%, as well as both individuals and SNPs with missing rates larger than 3%. This results in 645,568 SNPs with 487,048 individuals.

Details concerning UKB imputation are provided in [1]. UKB use the HRC reference dataset and a custom modification of "IMPUTE2" which employs a hidden-Markov-model. The imputed genotypes used for this work number 486,443 individuals. No further filtering was done.

#### 1.3 Phenotype Definitions and Predictor Construction

Phenotypes used in this paper are similar to those used in references [2–5]. Specifically, continuous phenotypes (height, bone mineral density and blood markers) are first z-scored by sex and corrected by a linear regression on year-of-birth. Case-control phenotypes are identified through ICD-10 codes, self-report data and hospitalization codes, please see [4] for specific codes used to identify cases.

Predictor construction is described extensively in references [2–5]. In summary: predictors are constructed using the implementation of LASSO regression (Least Absolute Shrinkage and Selection Operator) found in Scikit Learn for Python 3 [6] for all predictors except for Alzheimer's, Inflammatory Bowel Disease, Ischemic Stroke and Schizophrenia. All LASSO-constructed-predictors were trained and tested on self-reported European individuals in the UK Biobank. Specifically, the set of 40,000 individual who are in sibling pairs was set aside as a final testing set while the remaining 418,000 self-reported European individuals were used for training and validation. Alzheimer's [7], Inflammatory Bowel Disease [8], Ischemic Stroke [9] were trained using publicly available GWAS and PRS-CS with 1000 genomes LD information [10]. The Schizophrenia predictor used was a sparsified version of the publicly available predictor from the Pyschiatric Genomics Consortium [11].

### 2 deCODE comparison

The deCODE results are obtained from research published in 2019 [12]. See, specifically, supplementary material in the files: `aau1043_datas3.gz`, which contain the average paternal/maternal genetic maps and the cumulative centiMorgan location of the endpoint for each interval in GRCh38.

The additional figures here give cumulative results and recombination event density for each chromosome, averaged over 21k European ancestry siblings. We note three interesting anomalies on chromosomes 2, 15, 21 in which regions of high recombination activity are found, not present in the Icelandic data. We intend to investigate these results further in future work. It is possible that the anomalies result from specific mutations in those regions that increase recombination probability, and are more common in the UK population than in Iceland.

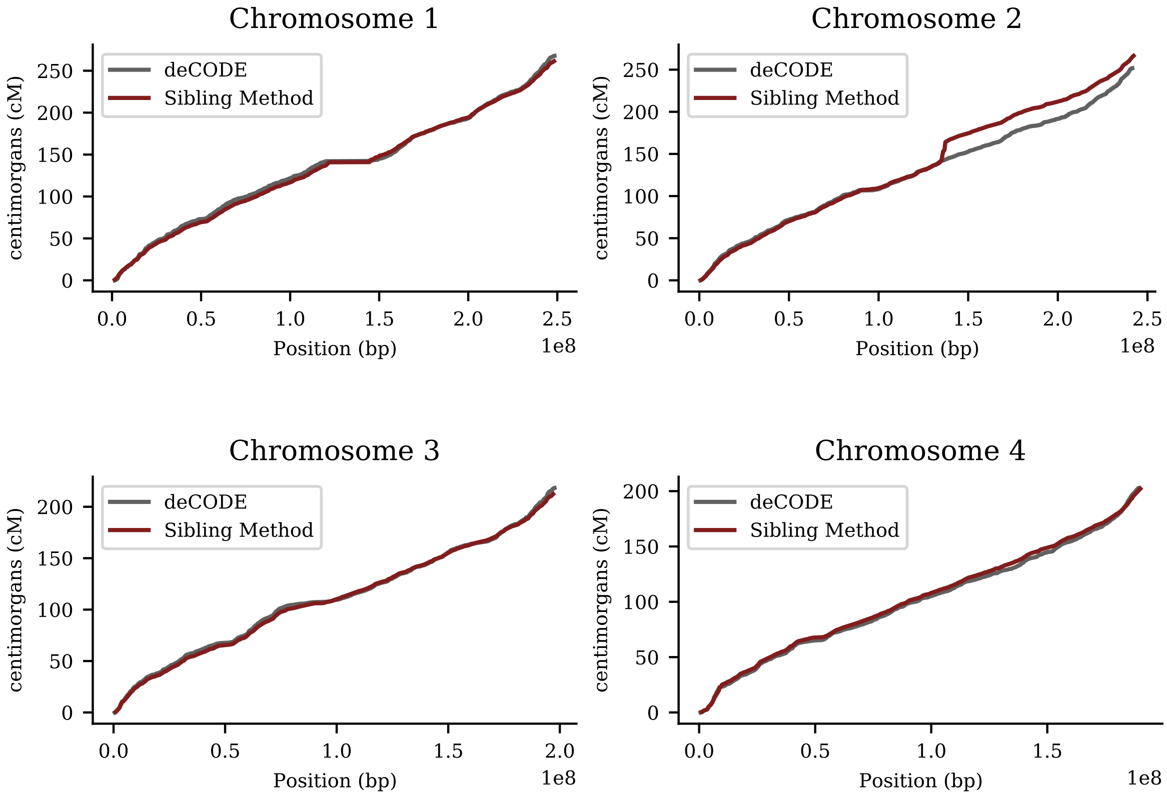


Figure 1: Cumulative recombination length of chromosomes 1-4 (i.e., total Morgan length). Red line is sibling-pair method applied to 21k European ancestry sibs from UK Biobank. Gray line is from deCODE (Science 2019) result using population of Iceland.

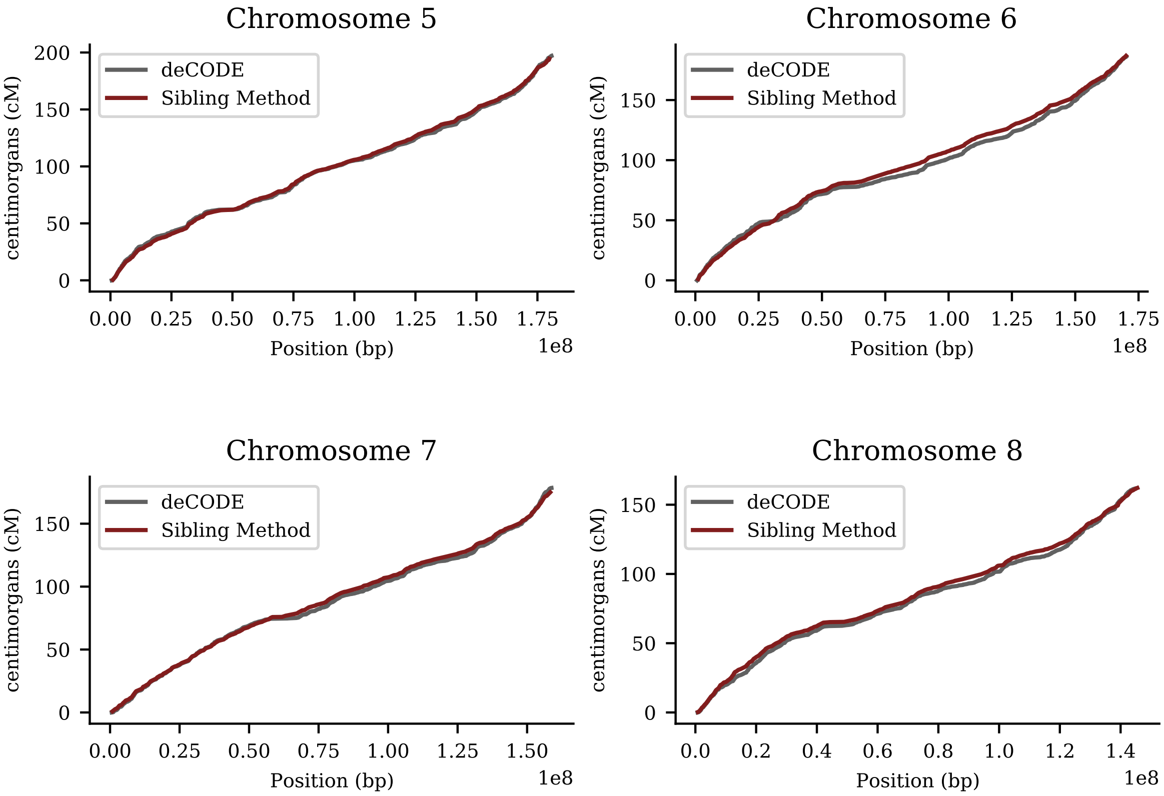


Figure 2: Cumulative recombination length of chromosomes 5-8 (i.e., total Morgan length). Red line is sibling-pair method applied to 21k European ancestry sibs from UK Biobank. Gray line is from deCODE (Science 2019) result using population of Iceland.

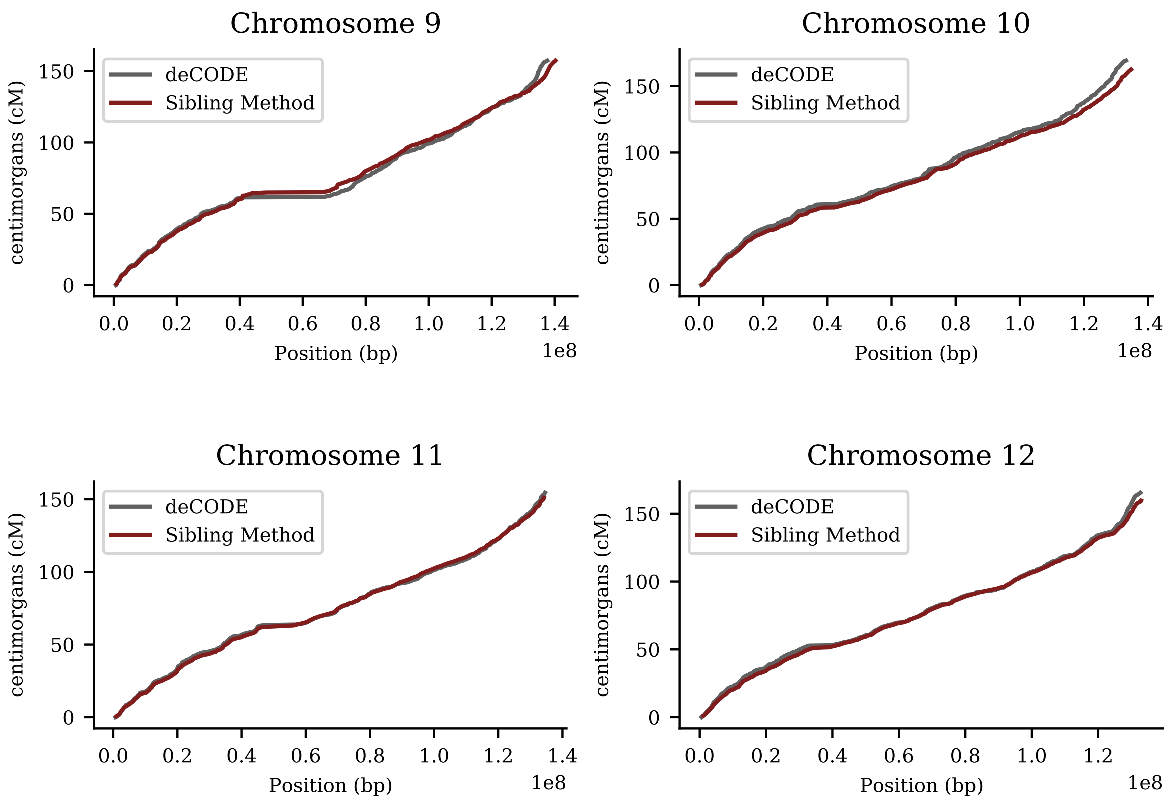


Figure 3: Cumulative recombination length of chromosomes 9-12 (i.e., total Morgan length). Red line is sibling-pair method applied to 21k European ancestry sibs from UK Biobank. Gray line is from deCODE (Science 2019) result using population of Iceland.

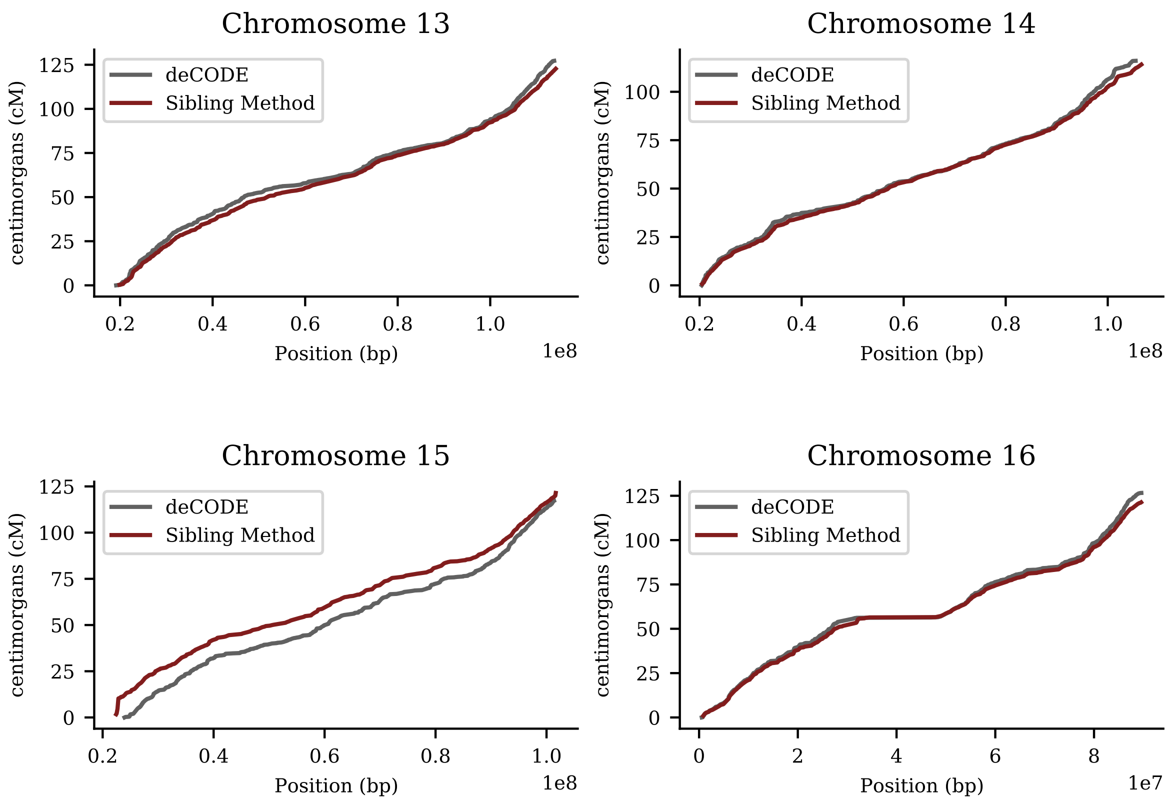


Figure 4: Cumulative recombination length of chromosomes 13-16 (i.e., total Morgan length). Red line is sibling-pair method applied to 21k European ancestry sibs from UK Biobank. Gray line is from deCODE (Science 2019) result using population of Iceland.

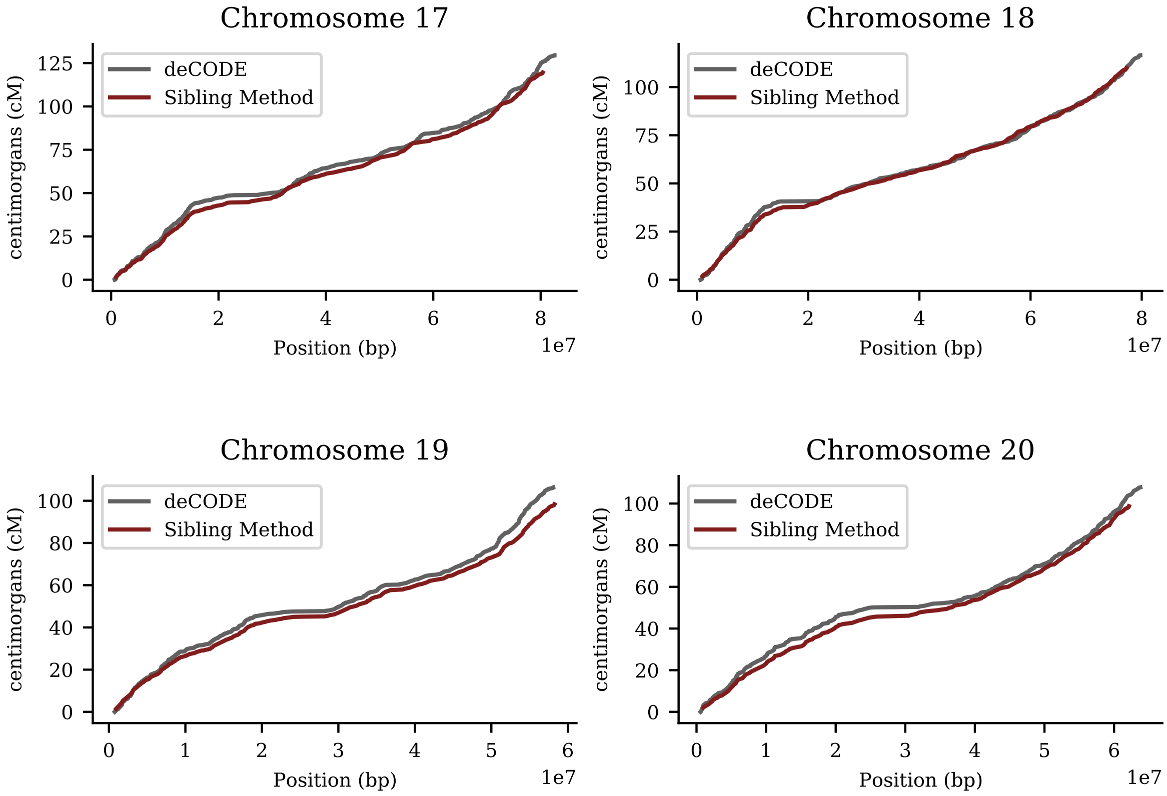


Figure 5: Cumulative recombination length of chromosomes 17-20 (i.e., total Morgan length). Red line is sibling-pair method applied to 21k European ancestry sibs from UK Biobank. Gray line is from deCODE (Science 2019) result using population of Iceland.

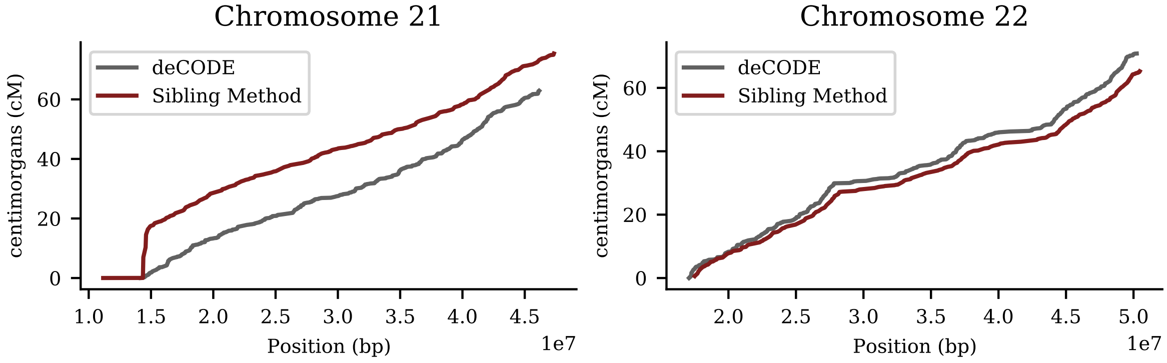


Figure 6: Cumulative recombination length of chromosomes 21,22 (i.e., total Morgan length). Red line is sibling-pair method applied to 21k European ancestry siblings from UK Biobank. Gray line is from deCODE (Science 2019) result using population of Iceland.

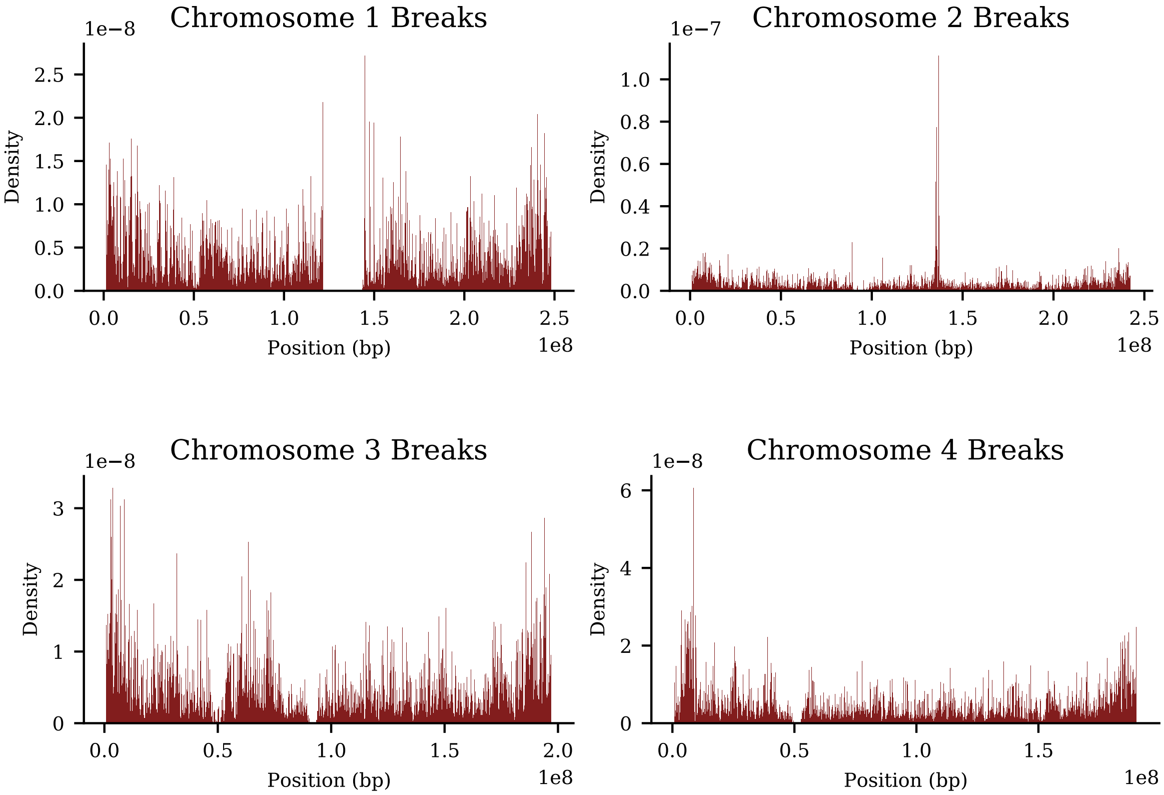


Figure 7: Density of recombination breaks on chromosome 1-4 from sibling-pair method, computed using 21k European sibling pairs.

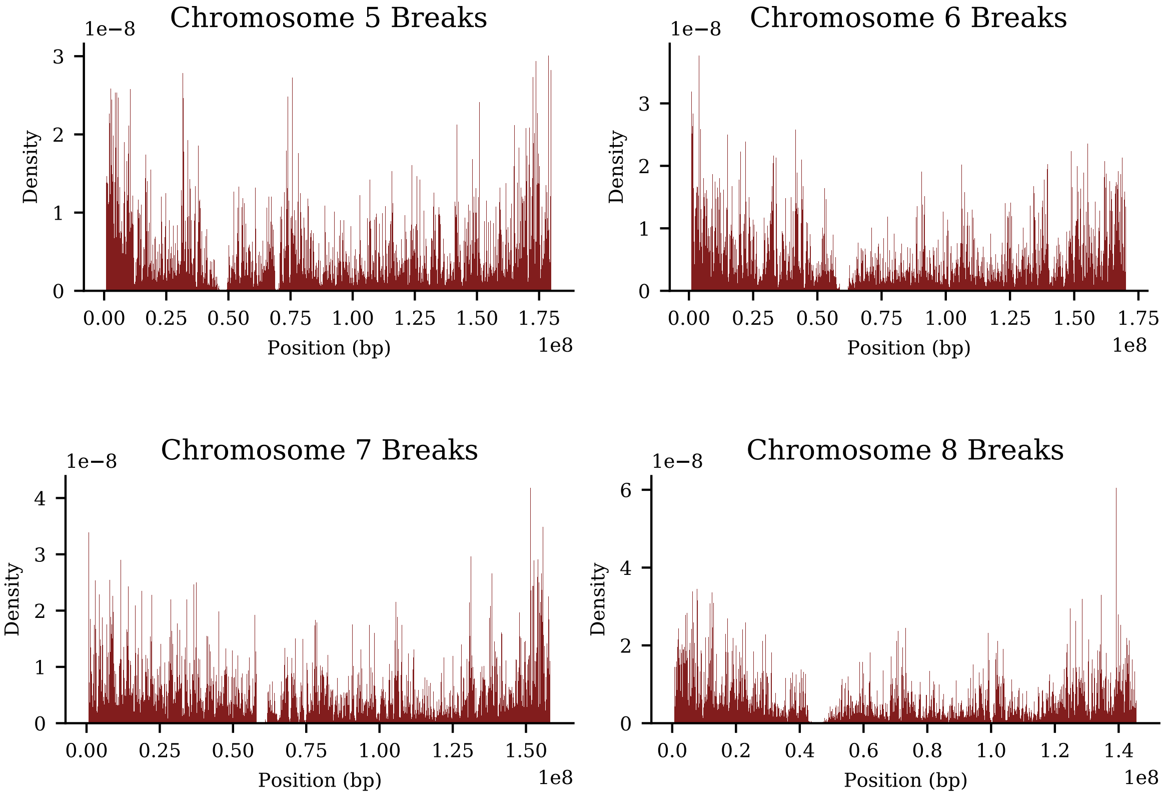


Figure 8: Density of recombination breaks on chromosome 5-8 from sibling-pair method, computed using 21k European sibling pairs.

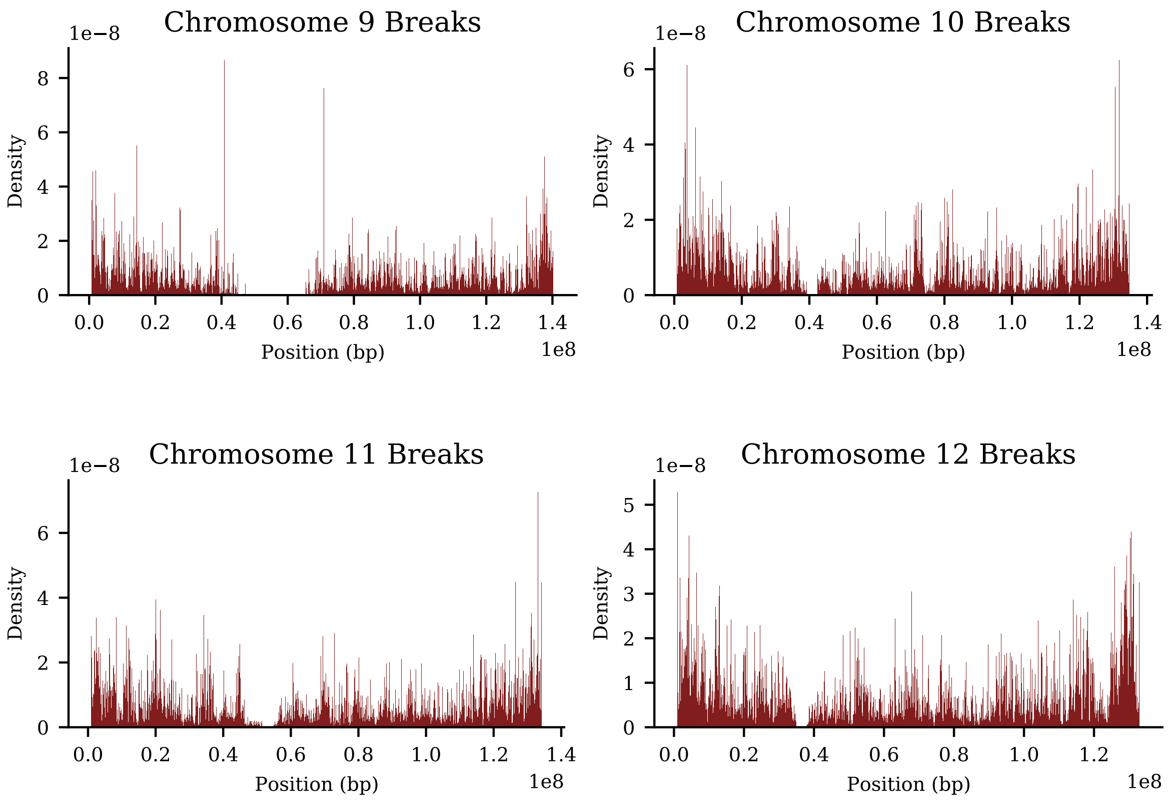


Figure 9: Density of recombination breaks on chromosome 9-12 from sibling-pair method, computed using 21k European sibling pairs.

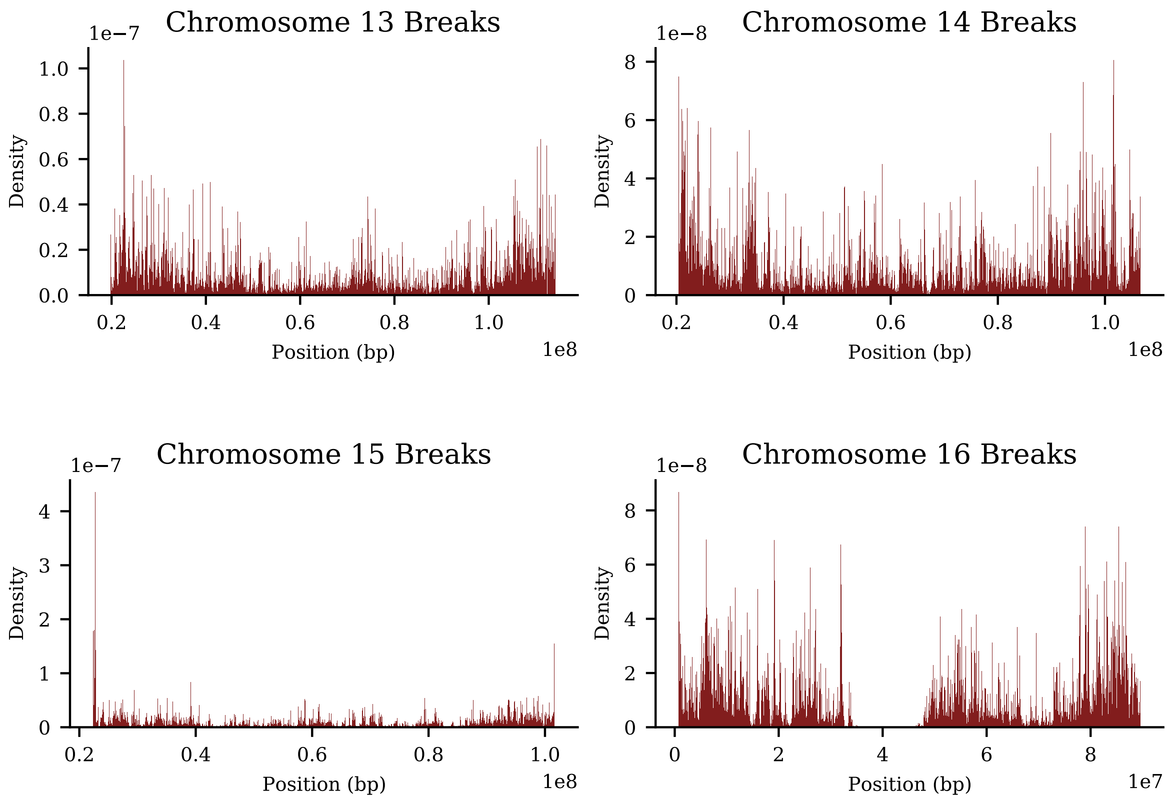


Figure 10: Density of recombination breaks on chromosome 13-16 from sibling-pair method, computed using 21k European sibling pairs.

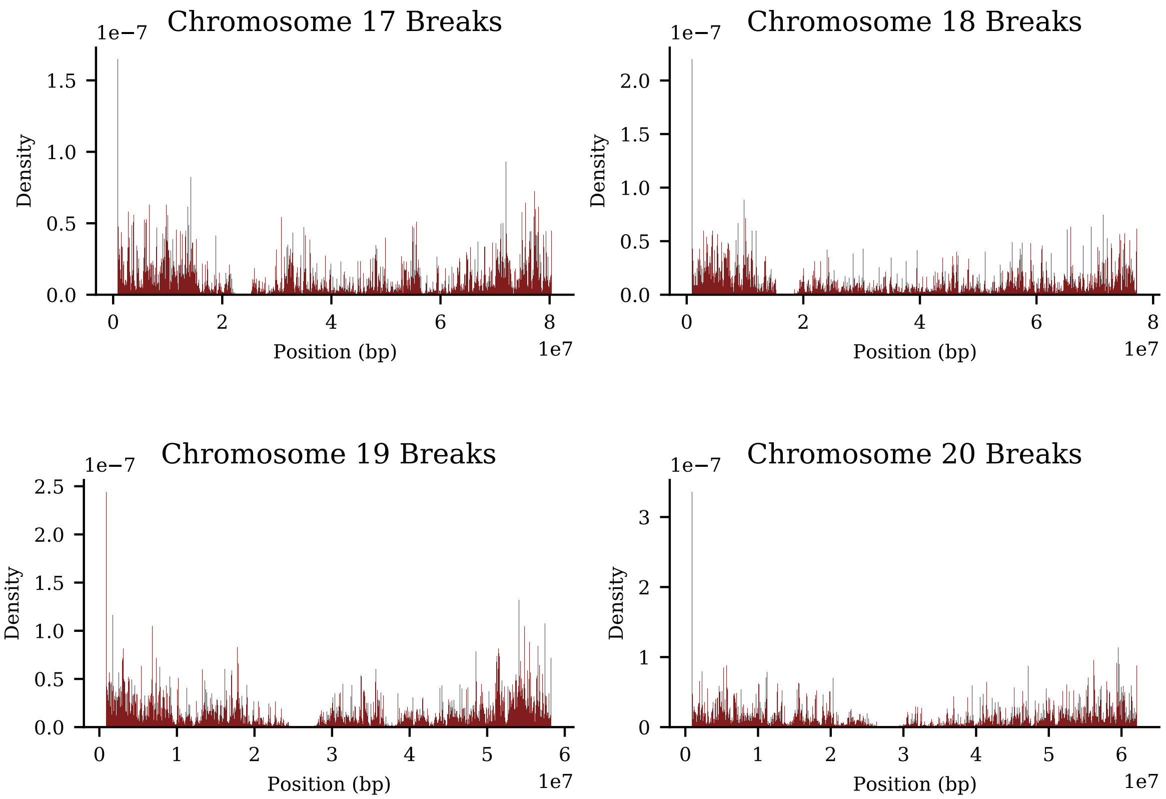


Figure 11: Density of recombination breaks on chromosome 17-20 from sibling-pair method, computed using 21k European sibling pairs.

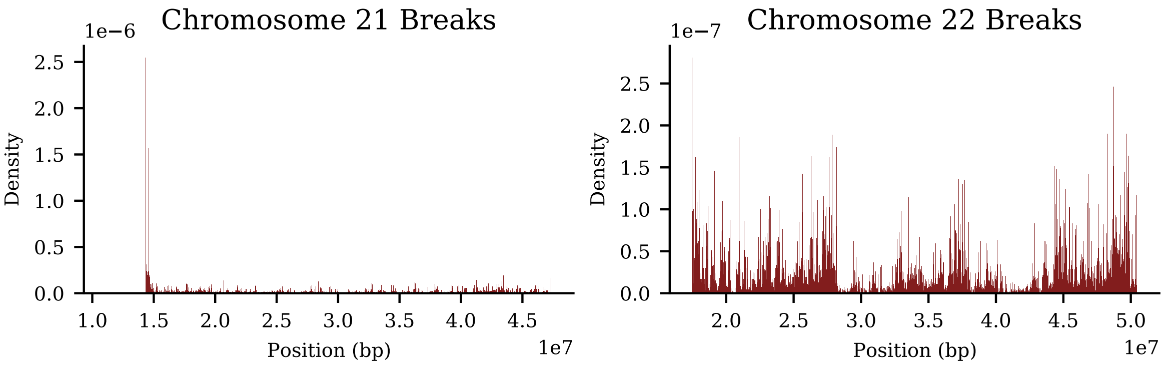


Figure 12: Density of recombination breaks on chromosome 21,22 from sibling-pair method, computed using 21k European sibling pairs.
